# Molecular age of Barrett’s esophagus indicates onset often occurs decades before diagnosis

**DOI:** 10.64898/2026.11.13.26367463

**Authors:** Talita Oliveira, Samuel R. Reynolds, Enya Scanlon, Victoria Child, Damian T. McManus, Brian Johnston, Zhijian Hu, Sarah Killcoyne, Barbara Nutzinger, Ginny Devonshire, Rebecca C. Fitzgerald, Richard C. Turkington, Amy Jayne McKnight, Helen G. Coleman, Kit Curtius

## Abstract

**Background:** Timely identification of Barrett’s esophagus (BE) is crucial for early esophageal adenocarcinoma (EAC) detection. Screening is recommended from age 50 in individuals with additional risk factors, but biological BE onset age remains unknown.

**Objective:** Estimate biological BE onset age and duration before clinical diagnosis (dwell time).

**Design:** We applied a BE epigenetic clock to 61 samples from 41 patients in a population-representative Northern Ireland cohort and 164 samples from 140 patients in an independent UK cohort. BE dwell times were compared between patients with concurrent dysplasia/EAC and age- and sex-matched non-dysplastic BE controls in these cross-sectional studies. In a longitudinal cohort, pre-progression biopsies from 31 future high-grade dysplasia (HGD)/EAC progressors were compared with 35 non-progressors.

**Results:** Median estimated BE onset ages in the cross-sectional cohorts were 29.5 (IQR=22.5-45.8) and 29.3 (IQR=20.4-39.0) years, respectively. These estimates were further supported by independent epidemiologic modeling of age-specific BE prevalence data from more than 70,000 BE cases. Estimated dwell times were longer in patients with concurrent dysplasia (median difference 14.3 years; p=0.0042) or EAC (12.5 years; p=0.0002) than matched non-dysplastic BE controls. Dwell time exceeding 19.8 years was associated with higher odds of HGD/EAC progression (OR=12.1, 95% CI 3.7-47.5; p=9.96 × 10⁻⁵).

**Conclusion:** BE may arise decades before diagnosis. Although these findings do not justify lowering screening age, they suggest one-time screening from age 50 years may detect existing high-risk BE. Further modeling is warranted to evaluate implications for population-based screening, alongside prospective validation of epigenetic tissue age for progression risk stratification.

**Key Messages:** *What is already known on this topic:* Increasing patient age is a major clinical risk factor for both esophageal adenocarcinoma (EAC) and its precursor lesion, Barrett’s esophagus (BE), but the biological age or dwell time of BE before diagnosis cannot be directly measured clinically, limiting our understanding of BE onset timing and past disease evolution.

*What this study adds:* Using a methylation-based molecular clock model applied to 294 tissue samples, we estimated that the median age of onset of BE is approximately 30 years old in 3 independent UK cohorts with BE. We found substantial interpatient variability in estimated BE onset age, significantly older BE tissue age in patients with adjacent EAC or dysplasia, and 12-fold higher odds of HGD/EAC progression for patients with older BE tissue age in a prospectively followed cohort.

*How this study might affect research, practice or policy:* These findings suggest that BE may arise decades before clinical diagnosis, providing a prolonged window for EAC prevention and supporting further investigation of one-time screening from age 50 to identify existing high-risk BE, rather than broadly lowering screening ages. Molecular estimates of BE tissue age may improve computational models of BE natural history and inform future biomarkers for neoplastic progression risk stratification, warranting further modeling and prospective validation to assess their potential utility in personalized screening and surveillance strategies.

## Introduction

The incidence of esophageal adenocarcinoma (EAC) and its precursor, Barrett’s esophagus (BE), increased from 1975 through the early 2000s^1,2^ before plateauing in the last decade^3,4^. Although only a small proportion of BE patients develop high-grade dysplasia (HGD) or EAC (∼0.2-0.4% annually^5,6^), identifying those at highest risk remains a major clinical challenge. Established risk factors of EAC progression include high body mass index (BMI), male sex, tobacco smoking, older age, ulceration within the BE segment, long BE segment length, and presence of dysplasia^7–9^. However, histopathologic detection of dysplasia, which minimally stratifies patients for a smaller endoscopic follow-up window,^10–12^ is the only indicator of progression that is widely used, despite its limited positive predictive value^13^ and substantial inter-observer variability^14^. Moreover, approximately 10% of BE patients develop HGD or EAC without any previous endoscopic indication such as low-grade dysplasia (LGD)^15^, highlighting the need for molecular biomarkers to improve risk stratification and enable more personalized surveillance rather than current ‘one-size-fits-all’ strategies.

Due to the likelihood that nearly all EAC arises in BE^16–18^, an important strategy for reducing EAC mortality is to identify BE patients before neoplastic progression and detecting HGD or early EAC during surveillance when treatment is most effective^19–22^. Current guidelines recommend screening in selected high-risk individuals with multiple risk factors, including age ≥50 years, male sex, white race, history of smoking, chronic gastroesophageal reflux disease (GERD), obesity, or family history of BE or EAC^10,12,23,24^. However, recent studies suggest that BE diagnoses rates are decreasing in older populations while increasing in younger populations, particularly in those aged 30-39 years^25,26^. Understanding when BE first develops and how long it persists before clinical detection may therefore help inform future screening strategies.

Although older chronological age is associated with increased EAC progression risk (60-69-year-old vs <50-year-old HR=2.43; 95% CI 1.43-4.13), substantial inter-patient variability^5,27^ may reflect differences in the age at which BE first develops^28^. Indeed, BE duration, or dwell time (patient age minus BE onset age), may be more biologically relevant to progression risk than chronological age alone, reflecting the progressive accumulation of somatic mutations, including TP53 alterations^29^, copy number changes^30^ and epigenetic alterations over time^31,32^. DNA methylation-based measures of biological aging have been associated with increased risk of neurodegenerative disease^33^, cardiovascular disease^34^, cancer^35^, and mortality^36,37^. However, BE onset age has historically been difficult to estimate because metaplastic tissue may persist asymptomatically for decades before diagnosis.

We previously developed a Bayesian molecular clock that uses age-associated DNA methylation drift at selected CpG sites to estimate patient-specific BE onset age and tissue dwell time^28^. Briefly, certain CpG dinucleotides are susceptible to methylation changes over time (clock*-* CpGs)^38,39^ and the incidence and accumulation of *de novo* methylation in some specific CpG sites, known as epigenetic drift, can be used to determine accelerated aging rates in BE tissue versus normal esophagus. Here, we applied this molecular clock to three UK study cohorts to characterize BE onset ages and dwell times, including validation against normal gastroesophageal tissues and independent epidemiologic modeling. We further compared BE tissue ages between age-matched patients with non-dysplastic BE and those with prevalent HGD/EAC and evaluated whether tissue age at initial BE diagnosis was associated with subsequent neoplastic progression in a matched longitudinal case-control cohort. Together, these analyses aimed to clarify the natural history of BE and assess the potential of molecular tissue age as a biomarker for personalized surveillance.

## Materials and Methods

### Study populations and DNA methylation profiling

We analyzed BE and non-BE gastroesophageal tissue samples from three identified UK study populations: the Northern Ireland Barrett’s oesophagus Register (NIBR) cross-sectional cohort (n=50 patients), the Cambridge Oesophageal Cancer Clinical and Molecular Stratification (OCCAMS)/Biomarker/BEST2 cohort^40,41^ (n=140 patients), and a nested NIBR case-control longitudinal cohort (NIBR-Progressor) of 42 HGD/EAC progressors and 42 matched non-progressors (n=84 patients). Tissue samples (formalin-fixed, paraffin-embedded [FFPE] from NIBR cohorts and fresh-frozen from Cambridge cohort) underwent expert pathological review. Cohort characteristics and selection criteria are detailed in the **Supplementary Information**.

After excluding samples that failed quality control (QC) and/or had missing demographic information on age at sampling (***Supplementary Figure S1***), the NIBR cohort included 35 patients with non-dysplastic BE (NDBE) and 6 with dysplastic BE. Patients with NDBE contributed biopsies from 1-4 time points, collected at incident endoscopy and during follow-up (≥6 months post-diagnosis), whereas patients with prevalent HGD contributed biopsies from a single time point at BE diagnosis. The Cambridge cohort included 140 patients, each contributing data from a single time sampling point at or after BE diagnosis: 22 with NDBE, 53 with adjacent EAC, 13 with adjacent HGD or intramucosal cancer (IMC), 19 with adjacent indefinite for dysplasia (IND) or low-grade dysplasia (LGD), 1 with adjacent crypt dysplasia, and 32 with unknown adjacent tissue dysplasia grade. In the NIBR-Progressor cohort, NDBE biopsies were collected at the time of BE diagnosis. In total, 291 samples were used in the downstream methylation analyses across the three cohorts: 61 from NIBR, 164 from Cambridge, and 66 from NIBR-Progressor.

Patient demographics and clinical characteristics are provided in ***Table 1*** (see ***Supplementary Figure S2*** for detailed study design). Genome-wide DNA methylation for each tissue sample was profiled using Illumina MethylationEPIC arrays (NIBR cross-sectional and Cambridge cohorts) or the Twist Human Methylome Panel (NIBR-Progressor cohort). Platform-specific preprocessing, normalization, and quality control procedures are described in the **Supplementary Methods.**

**Table 1.** Demographic and clinical characteristics of patients in the Northern Ireland Barrett’s oesophagus Register (NIBR), Cambridge, and NIBR-Progressor cohorts following quality control filtering. In the Cambridge cohort, age of BE diagnosis was available for 17 patients. Race/ethnicity, family history, and year of sampling was available for all non-EAC patients in the Cambridge studies (n=93).

| **Data Type** | **Variable** | **NIBR** | **Cambridge** | **NIBR-Progressor** |
|---|---|---|---|---|
| Cases | Number of patients | 41 | 140 | 66 |
|  | Number of samples | 61 | 164 | 66 |
| Highest pathology grade diagnosed for the patient at time of tissue sample | Non-dysplastic BE | 55 (90.2%) | 27 (16.5%) | 66 (100%) |
|  | IND/LGD | 0 | 19 (11.6%) | 0 |
|  | HGD/IMC | 6 (9.8%) | 17 (10.5%) | 0 |
|  | Crypt dysplasia | 0 | 1 (0.55%) | 0 |
|  | EAC | 0 | 66 (40.2%) | 0 |
|  | Unknown | 0 | 34 (20.7%) | 0 |
| Sample tissue type | Barrett's esophagus | 61 (100%) | 115 (70.1%) | 66 (100%) |
|  | Gastric cardia | 0 | 25 (15.3%) | 0 |
|  | Squamous esophagus | 0 | 24 (14.6%) | 0 |
| Patient clinical information | Sex (% male) | 65.6% | 84.1% | Progressor (n=31):<br>84.4%<br>Non-progressor (n=35):<br>82.4% |
|  | Race/ethnicity<br>White: British<br>White: Other<br>N/A | 41 | (n=93 non-EAC)<br>32<br>4<br>57 | 66 |
|  | Family history<br>of GERD<br>of BE<br>of EAC<br>of other cancers<br>None<br>N/A | 41 | (n=93 non-EAC)<br>10<br>0<br>0<br>0<br>6<br>77 | 66 |
|  | Median age at BE diagnosis (years) | 56 (IQR = 45-66) | 63.21 (IQR = 56.2-77.3)<br>(n=17) | Progressor (n=31):<br>59 (IQR = 54-65.5)<br>Non-progressor (n=35):<br>59 (IQR = 51- 65) |
|  | Median age at sampling (years) | 56 (IQR = 46-66) | 68.8 (IQR = 62-76) | Progressor (n= 31):<br>59 (IQR = 54-65.5)<br>Non-progressor (n=35):<br>59 (IQR = 51- 65) |
|  | Median year of sampling | 1999 (IQR = 1998-2000) | 2014 (IQR = 2011-2015)<br>(n=93 non-EAC) | Progressor (n=31):<br>2009 (IQR = 2004-2011)<br>Non-progressor (n=35):<br>2008 (IQR = 2022-2011) |

\* IQR = Interquartile range, N/A = Not available

### BE onset age and tissue age estimation

We applied a previously developed Bayesian epigenetic clock model based on methylation drift at BE-specific CpG sites to estimate patient-specific BE onset ages using Markov chain Monte Carlo sampling^28^. BE tissue age (dwell time) was calculated as patient age at biopsy minus estimated onset age. Sensitivity analyses assessed robustness of onset age estimates to variation in CpG data availability across samples, and estimates were compared with those from two independently developed epigenetic clock models^42,43^ (see **Supplementary Methods**). Mann-Whitney U tests and logistic regression evaluated group differences and associations with neoplastic progression.

### Epidemiologic modeling

Independently, we inferred the population distribution of BE onset ages by fitting a scaled gamma cumulative distribution function to published age-specific BE prevalence data from three U.S. study populations (n=77,832 total BE patients)^44–46^. Simulated onset ages conditional on observed endoscopy ages were used to estimate cohort-specific distributions, with 95% simulation intervals obtained from 10,000 Monte Carlo replicates (see **Supplementary Methods** for details).

## Results

We analyzed BE biopsies obtained from patients at a median age of 56 years (IQR: 45-66) in the NIBR cohort and 59 years (IQR: 53.3-65) in the NIBR-Progressor cohort, corresponding to their respective ages at BE diagnosis (***Table 1***). Most patients were male, and male patients had a median age at BE diagnosis of 50 years (IQR = 42-60) in NIBR and 58 years (IQR = 53.5-63.5) in NIBR-Progressor cohorts. Female patients had a median diagnosis age of 70 years (IQR= 56-73) in the NIBR and 66 years (IQR 57.5-67) in NIBR-Progressor cohorts. Patients with HGD in the NIBR cohort had a median BE diagnosis age of 67.5 years (IQR = 56-73) at the time of prevalent HGD diagnosis. Male and female patients had a median age of BE sampling of 68.0 years (IQR = 61.8-75.0) and 73.5 years (IQR = 66.2-79.0) in the Cambridge cohort, respectively. Patient-specific race/ethnicity data is not available for patients in NIBR-derived study populations; however, the Northern Ireland population was 96.5% White in the 2021 Northern Ireland Census data.

For all patients, an NDBE sample was used for BE onset age estimation, and we used estimates for normal samples as BE clock validation. For NIBR and Cambridge cohorts (***Figure 1A***), beta-values at previously identified clock-like CpG sites specific to BE were obtained from methylation arrays for each sample, which confirmed greater epigenetic drift toward higher methylation in BE compared to normal samples and variable degree of drift across metaplastic samples (***Figure 1B***)^28^. Similarly, for the NIBR-Progressor cohort, beta-values at the previously identified clock-like CpG sites specific to BE were obtained from Twist platform data for each sample (***Figure 1C***). BE onset age for each sample was estimated using our BE epigenetic clock model^28^ (see **Materials and Methods**) and final estimated BE onset age results were obtained for 61, 147, and 66 samples from NIBR, Cambridge, and NIBR-Progressor respectively. Onset ages for 17 samples (14 normal squamous samples, 2 normal gastric cardia samples, and 1 BE sample) could not be estimated from the data due to lack of Bayesian inference convergence (details below).

**Figure 1.**
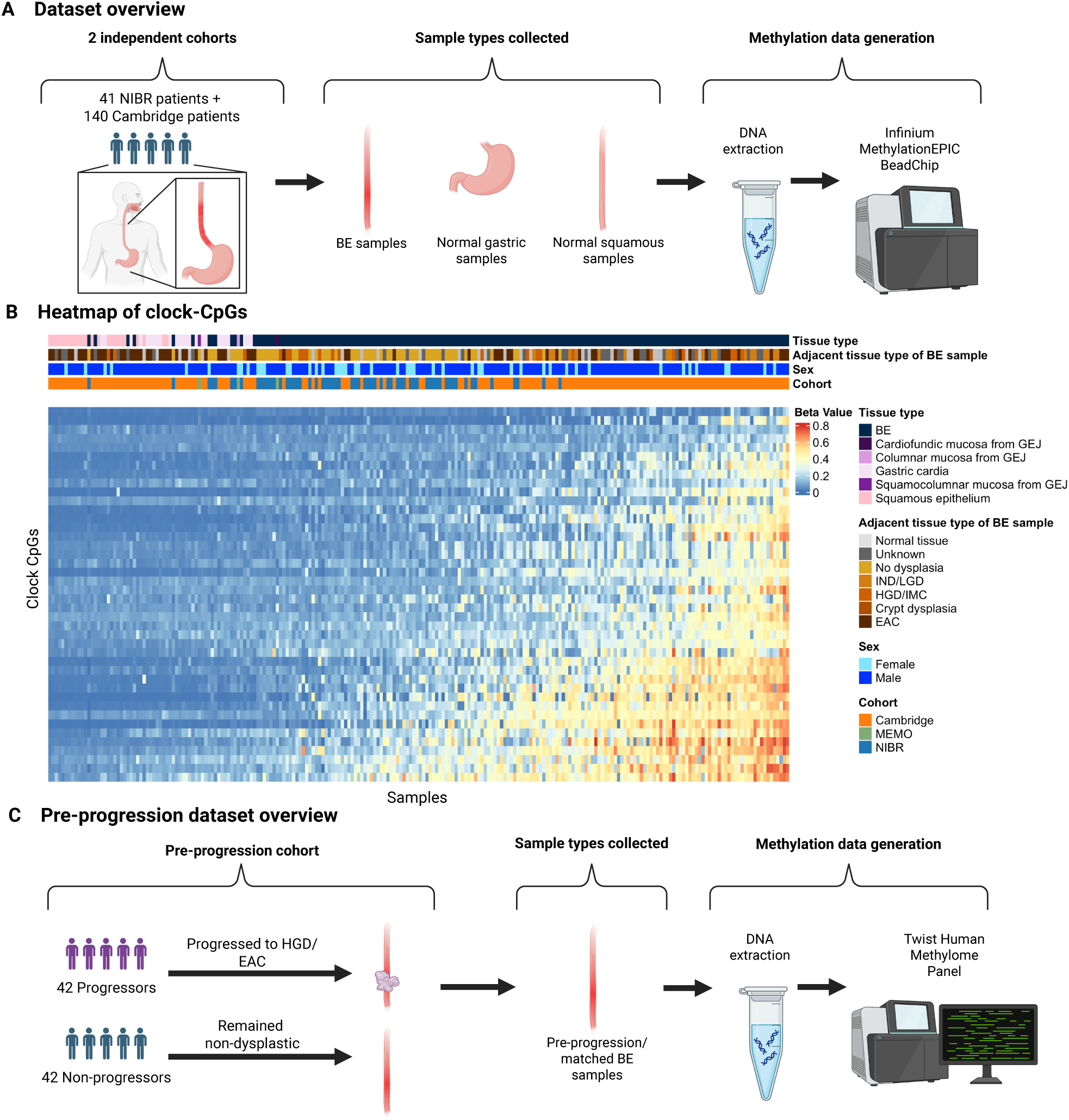
Study design. **A.** Overview of NIBR and Cambridge cohort datasets. **B.** Heatmap of epigenetic drift (beta value) at BE clock CpG sites in each sample (n=225, including data for 3 tissue samples from the gastroesophageal junction (GEJ) as a comparison), ordered by mean beta value across sites/samples in NIBR and Cambridge cohorts. Higher methylation at BE clock-associated CpG sites indicates advanced biological age of the tissue. **C.** Overview of nested case-control study design of the longitudinal NIBR-Progressor cohort, comparing non-dysplastic BE (NDBE) biopsies collected at BE diagnosis before progression to high-grade dysplasia (HGD) or esophageal adenocarcinoma (EAC) with biopsies from age-matched non-progressors.

### Epigenetic clock validation with normal tissue samples

The BE molecular clock uses epigenetic divergence from the normal epigenome (esophageal squamous or gastric cardia) to predict BE onset age. For two patients of the same chronological age at sampling, the patient whose BE developed earlier would have a younger estimated BE onset age and, consequently, a longer BE dwell time. In contrast, normal gastroesophageal epithelium is expected to have an onset age that is similar to the age of sampling (i.e., the clock model ‘ticks’ with age of the patient rather than BE age) and a dwell time of ∼ 0 based on how the BE clock was originally derived^28^. We found statistically significant different estimated dwell times between non-BE and BE tissues (gastric cardia [GC] vs. BE: p = 1.0e-12; squamous esophagus [SQE] vs. BE: p = 9.0e-8; gastroesophageal junction [GEJ] vs. BE: p = 0.018), with negligible median dwell times in normal tissues (GC: median = 1.8 years [IQR = 1.3-3.8]; SQE: median = 0.7 years [IQR = 0.6-0.8]; ***Supplementary Figure S3A, B***). 16 of the 49 normal samples (14 SQE and 2 GC, thus not shown in ***Supplementary Figure 3***) had onset estimates indiscernible from patient age (dwell time = 0) given our Markov Chain Monte Carlo prior constraints. In contrast, the median dwell time in BE tissue across all cohorts (NIBR, Cambridge, and NIBR-Progressor) was 30.6 years (IQR = 16.0-40.1). The clock therefore behaves as expected in cohorts independent of those used for model development^28^, supporting its validity and applicability.

### Similar distributions of BE onset age and dwell time estimates in two independent cohorts

Estimated BE onset ages are shown in ***Figure 2*** for the NIBR cohort at time of BE diagnosis and in ***Figure 3*** for the Cambridge cohort at time of BE tissue sampling. The median BE onset age in the NIBR cohort was 29.5 years (IQR = 22.5-45.8), which was significantly lower than the age of BE diagnosis (median = 50.0; IQR = 41.8-60.5; p = 0.0003, ***Figure 2B***). 87.5% had a BE onset age of less than 50 years old, with 12.5% estimated to have the onset of disease be between 10 and 20 years old, 41.6% between 20 and 30 years old, 4.2% between 30 and 40 years old, and 29.2% between 40 and 50 years old. The remaining 12.5% of patients had a BE onset age between 50 and 57.6 years old. The median BE dwell time for these patients was 18.1 years (IQR = 11.2-23.8).

**Figure 2.**
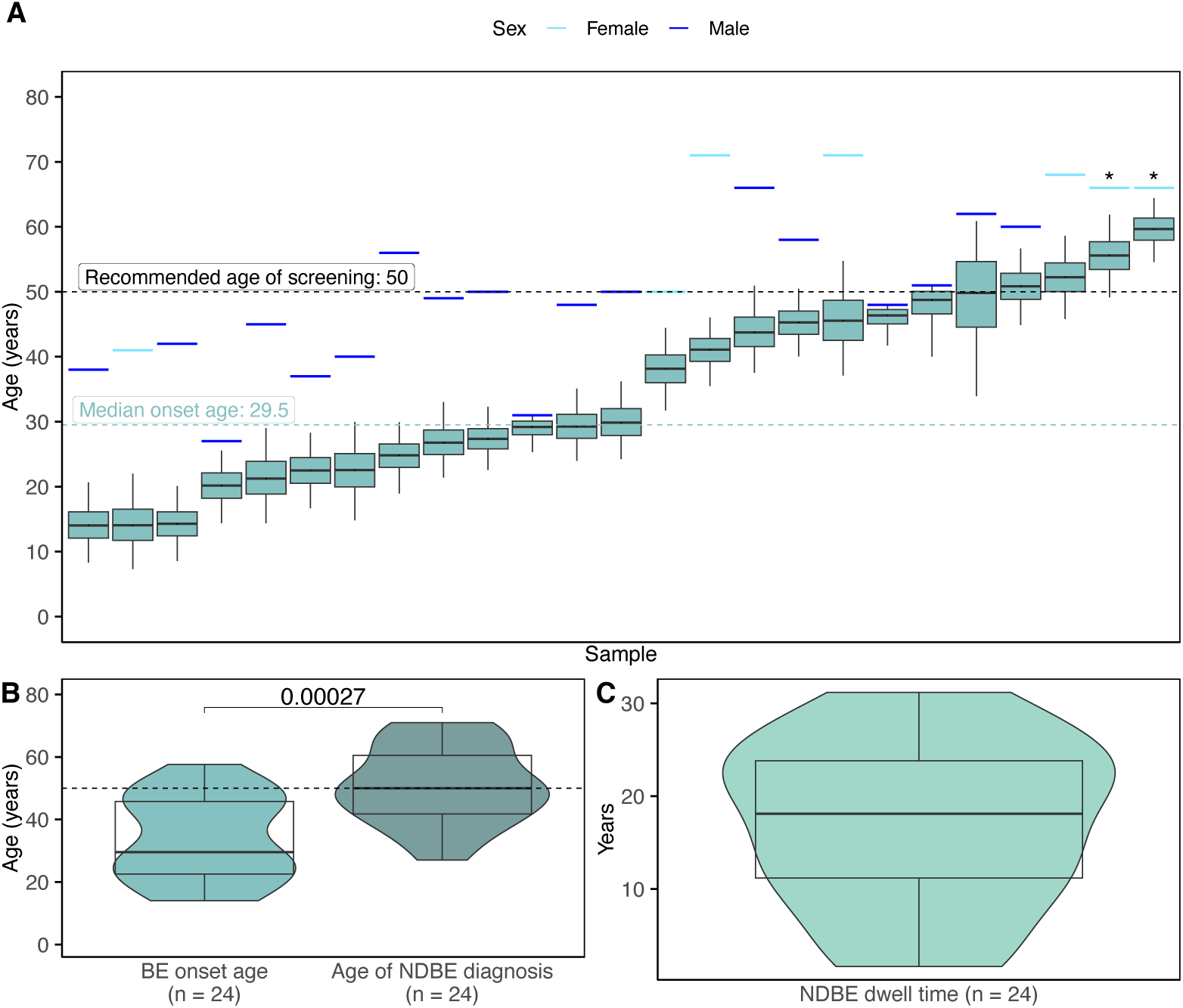
Estimated BE onset ages and dwell times for population-representative cohort. **A.** Patient-specific BE onset age estimates for the NIBR cohort at the first sampling timepoint (BE diagnosis). Horizontal bars indicate age at sampling for females (light blue) and males (dark blue), while boxplots represent the Bayesian posterior distributions of estimated BE onset age for each patient (see **Materials and Methods**). Patients are ordered by increasing median estimated onset age. Asterisks indicate 2 biopsy samples collected from the same patient. **B.** Violin plots comparing estimated BE onset age with age at clinical diagnosis. For patient with duplicate samples, the mean of the two estimates was used. **C.** Violin plot of corresponding estimated BE dwell times, calculated as age at sampling minus estimated BE onset age.

**Figure 3.**
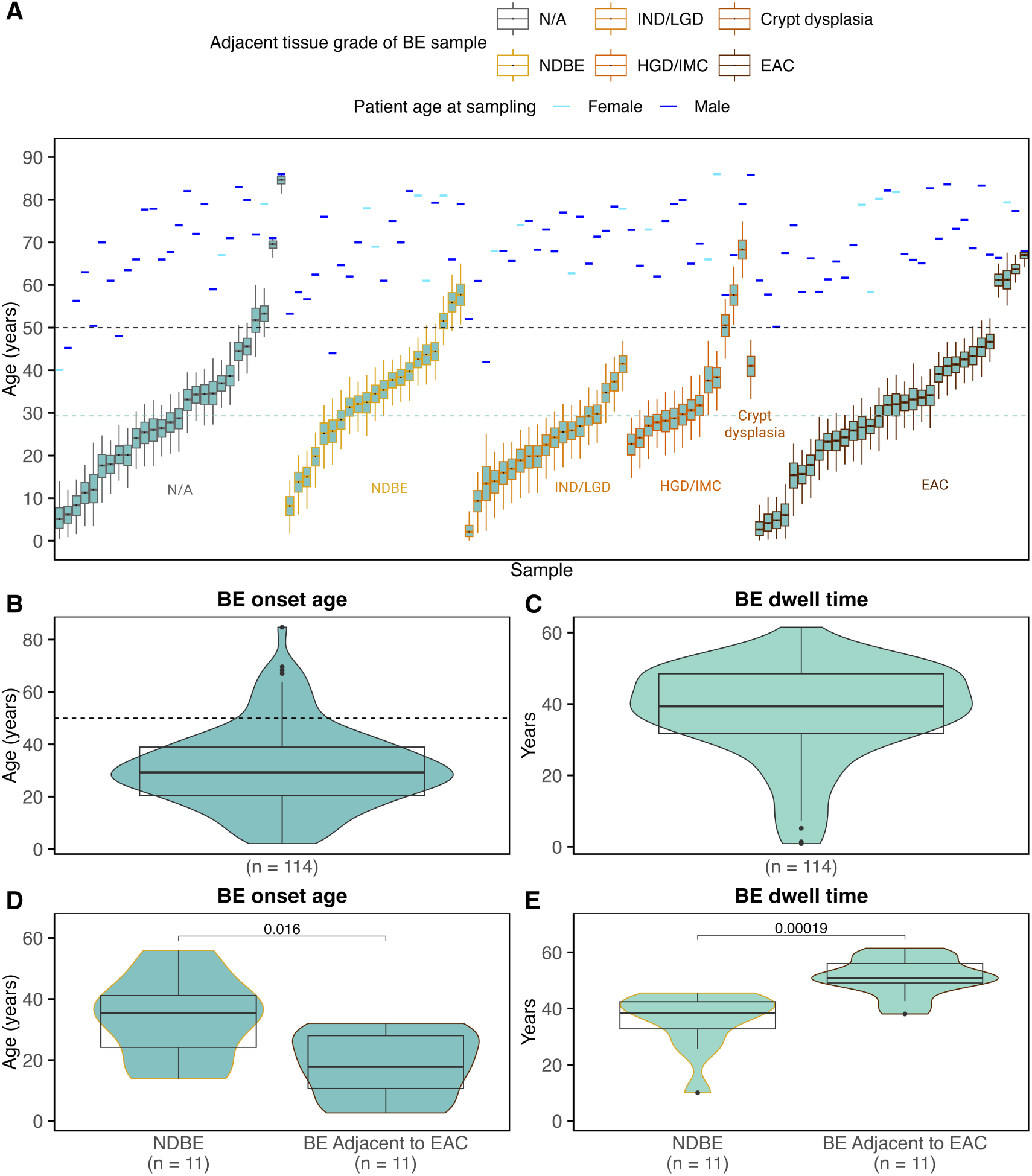
Estimated BE onset ages and dwell times in patients with and without adjacent neoplasia. **A.** Light blue (female) and dark blue (male) horizontal bars indicate chronological age at BE sampling, while boxplots represent the Bayesian posterior distributions of estimated BE onset age for each patient (see **Material and Methods**). Patients are grouped according to the highest grade of neoplasia adjacent to the BE sample used for tissue age estimation, where available. Black horizontal dotted lines indicate age 50, and the pale blue dotted line indicates the median estimated BE onset age (29.3 years) across the combined Cambridge cohort. **B.** Violin plots of median estimated BE onset ages derived from posterior distributions for patients shown in panel A. **C.** Violin plots of estimated BE dwell times, calculated as chronological age at biopsy minus median estimated BE onset age. **D.** Comparison of median estimated BE onset ages between matched non-dysplastic (NDBE) patients and EAC patients. **E.** Comparison of median estimated BE dwell times between matched NDBE and. EAC patients. Patients were matched by sex and chronological age at BE sampling (within 2 years) in panels D and E.

The median estimated BE onset age in the Cambridge cohort (29.3 years; IQR = 20.4–39.0) was similar to that of NIBR (p=0.34). 87.7% of the Cambridge patients had an onset age of less than 50 years old, with 8.8% estimated to have the onset of disease before the age of 10, 14.9% between 10 and 20 years old, 28.9% between 20 and 30 years old, 23.7% between 30 and 40 years old, and 11.4% between 40 and 50 years old. The remaining 12.3% of patients had a BE onset age between 50 and 84.7 years old. The median dwell time for these patients was 39.3 years (IQR = 31.8-48.5).

In a sensitivity analysis of patients with available symptom data, we found no statistically significant difference in estimated BE onset age between symptomatic and asymptomatic patients receiving acid-suppressive therapy (***Supplementary Figure S4***). Although females tended to have later estimated BE onset ages than males, neither BE onset age nor dwell time differed significantly between sexes after matching on chronological age at sampling (***Supplementary Figure S5***).

### Estimated BE onset age and dwell time indicate advanced BE tissue age in patients with EAC

Estimated BE onset and dwell time between patients with adjacent EAC versus those with NDBE were compared in ***Figure 3D*** and ***3E***, respectively. When matching for age at BE sampling and sex (***Supplementary Figure S6***), estimated BE onset age was significantly younger in patients with EAC (median 17.8 years, IQR = 10.7-28.0) than in those with NDBE at time of sampling (median = 35.4 years, IQR=24.1-41.2; p = 0.016). Additionally, the median estimated BE dwell time in those with NDBE was 38.4 years (IQR = 32.8-42.4), significantly shorter than the dwell times in those who progressed to EAC (median = 50.9 years, IQR = 49.1-56.0; p = 0.00019). Taken together, we estimated that BE patients with EAC had developed BE a median of 12.5 years earlier than matched BE patients who had not progressed to neoplasia.

### BE onset age generally consistent across serial time points

BE onset age and dwell time were also estimated for sequential biopsies taken from the same patient in the NIBR cohort over time. Some patients with sequential biopsies had only later timepoint samples analyzed after QC filtering and were included in this analysis. For indolent BE patients in NIBR, we analyzed data for 19 patients with 1 time point, 14 patients with 2 time points, 0 patients with 3 time points, and 2 patients with 4 time points. BE onset age estimates across timepoints were typically consistent and the median difference between BE onset age estimated from time points 1 and 2 in the 12 patients with available data was 0.4 years (pairwise Wilcoxon p=1, ***Supplementary Figure S7***). One potential reason for differences in predicted BE onset age is difference in location/level of biopsy sample within the BE segment over time, but we did not have accurate information on this variable in NIBR to test for any spatial trends.

### Validation of BE dwell times with independently developed epigenetic clocks

To further validate our model and compare its BE onset age estimates with independent measures of epigenetic aging in these two cohorts, we applied two orthogonal epigenetic aging clocks to our BE data. We first applied epiTOC1 which reports the average beta value across 385 CpGs associated with Polycomb target genes that are known to be unmethylated at birth and gradually drift toward hypermethylation throughout life^42^. Then we applied epiTOC2 which estimates the number of mitotic divisions that have occurred throughout life in that tissue^43^. Consistent with the substantial heterogeneity in BE dwell times estimated using our molecular clock (***Figure 2C*, *Figure 3C***), we observed substantial variability in epiTOC1 scores and epiTOC2 mitotic division rates (total number of mitotic divisions divided by chronological age at time of sampling) across BE tissue samples (***Figure 4 A,C***). For both epiTOC clock measures, we found the scores to be significantly higher in BE than the scores derived from the gastric cardia (GC), squamous esophagus (SQE), and gastroesophageal junction (GEJ) tissues (e.g., p = 5.9e-13 for epiTOC and p=2.6e-13 for epiTOC2 in the BE vs. GC comparison). Additionally, we found our dwell time estimates to be highly correlated with the epiTOC1 (R^2^ = 0.9, p < 2.2e-16) and epiTOC2 (R^2^ = 0.61, p < 2.2e-16) scores when applied to the same tissue samples (***Figure 4 B,D***).

**Figure 4.**
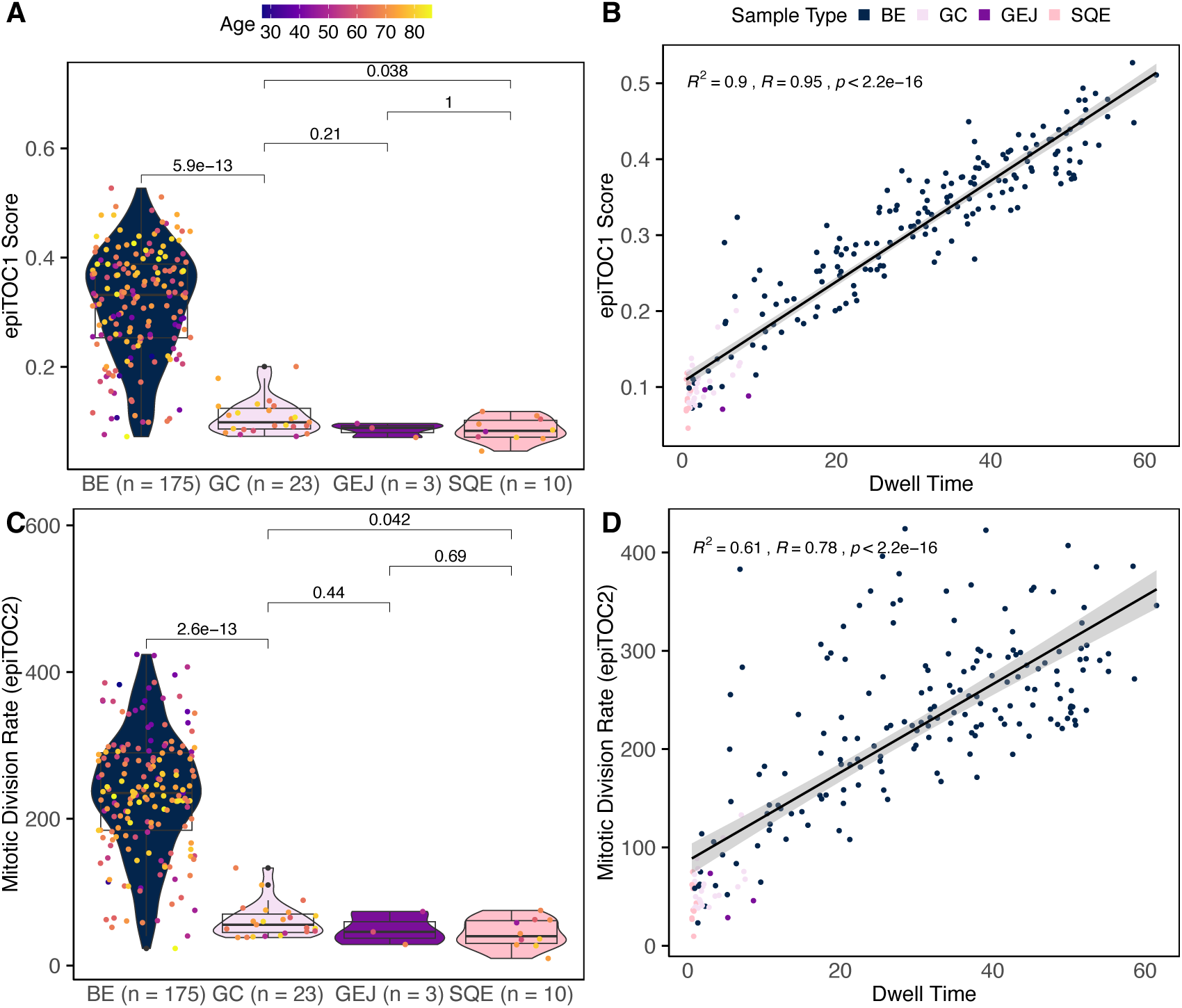
Validation of BE tissue age estimates with independent epigenetic clock models. **A.** epiTOC1 score across BE and non-BE tissues: gastric cardia (GC), gastroesophageal junction (GEJ), and squamous esophagus (SQE). **B.** Correlation between epiTOC1 score and estimated BE dwell time**. C.** epiTOC2 score across BE and non-BE tissues. **D.** Correlation between epiTOC2 score and estimated BE dwell time. Neither epiTOC1 nor epiTOC2 was developed or originally validated using BE tissue samples.

### Patients with adjacent dysplasia have biologically older BE than patients with indolent BE

To further investigate BE dwell times in relation to pre-EAC neoplastic risk, 13 BE patients with prevalent dysplasia (one of which had a duplicate biopsy) or IMC, were matched by age at sampling (within 2 years) and sex with 13 BE patients with NDBE in the NIBR and Cambridge cohorts. The 13 patients with NDBE were independent from those used in the comparison to EAC patients in ***Figure 3 D, E***. BE onset ages and dwell times for all patients were already estimated above from NDBE tissue (***Figures 2*, *3***). The median age at BE diagnosis for the dysplastic patients was 62 years old (IQR=54.0-66.0); the median onset age of BE adjacent to dysplasia was 27.8 years old (IQR=15.5.2-37.6); and the median dwell time was 34.2 years (IQR=28.4-40.3). The median age at diagnosis, onset age, and dwell time for the matched indolent BE cases were 62 years old (IQR=54.0-66.0), 45.6 years old (IQR=24.8-51.1) and 19.9 years (IQR=12.9-25.4), respectively (***Figure 5***). Consistent BE onset ages were found for one dysplastic patient (samples 2B and 2C in ***Figure 5A***) when analyzing methylation data separately from two BE biopsies taken from different esophageal quadrants. The dysplastic patients had significantly earlier BE onset ages (median = 27.8 years old, IQR = 15.5-37.6) than matched NDBE patients (median = 45.6 years old, IQR = 24.8-51.1) for a median difference in BE onset age of 17.8 years (p=0.044; ***Figure 5B***). Further, the median BE dwell times were significantly longer in the dysplastic patients (median = 34.2 years, IQR = 28.4-40.3) compared with the age and sex-matched non-dysplastic BE patients (median = 19.9 years, IQR = 12.9-25.4) for a median difference of 14.3 years (p=0.0042; ***Figure 5C***).

**Figure 5.**
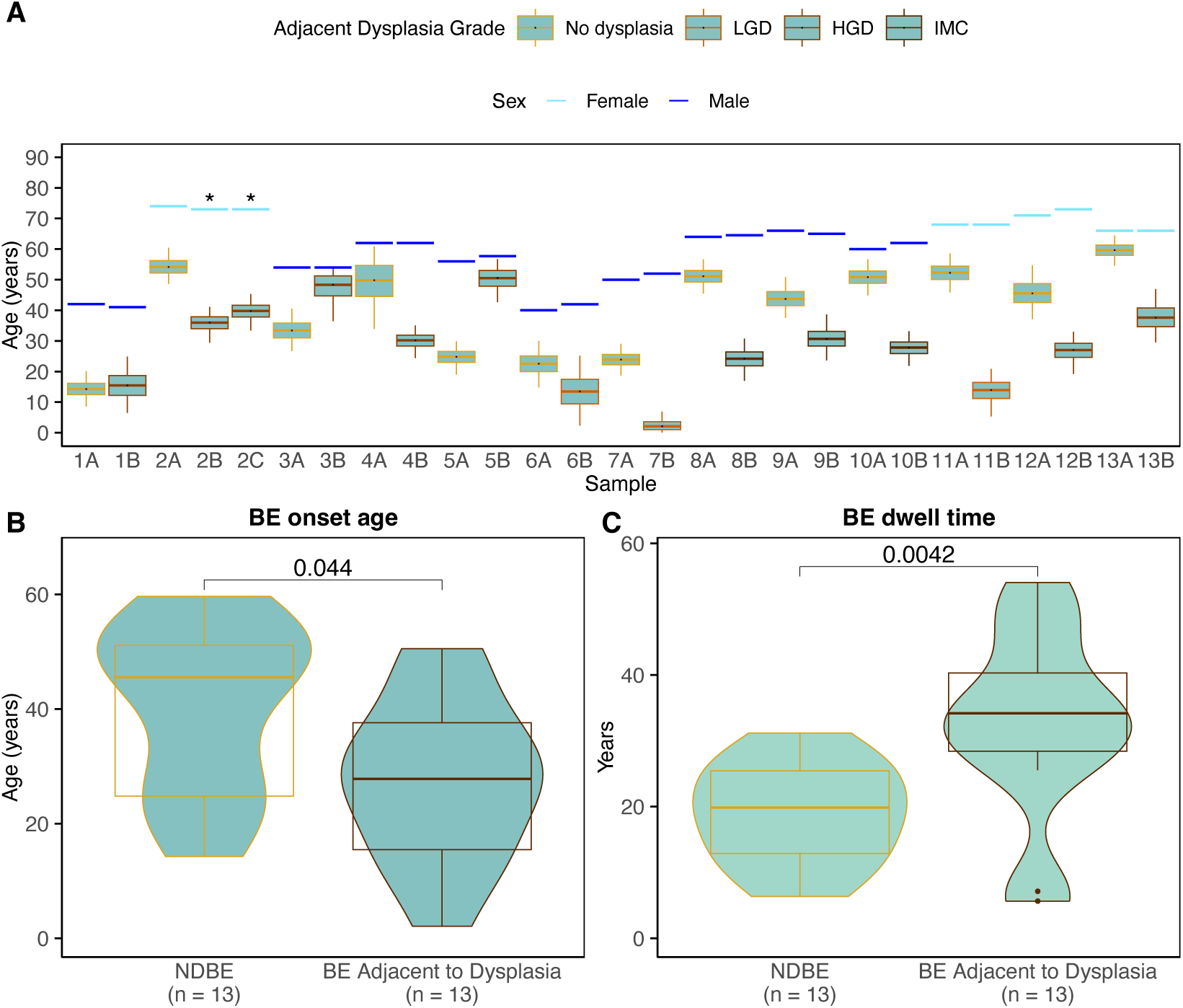
Comparison of estimated BE onset age and dwell time between patients with non-dysplastic BE (NDBE) and patients with BE adjacent to dysplasia. A. Light blue (female) and dark blue (male) horizontal bars indicate chronological age at BE sampling, and boxplots represent the Bayesian posterior distributions of estimated BE onset age for each patient (see Materials and Methods). Patients with NDBE were matched to patients with BE adjacent to dysplasia or intramucosal carcinoma (IMC) by sex and age at sampling (within 2 years). Pair 2 included duplicate BE samples adjacent to HGD (denoted by *), which were averaged in subsequent comparisons. B. Comparison of median estimated BE onset ages between matched groups. C. Comparison of median estimated BE dwell times between matched groups.

### Estimated BE dwell time is associated with subsequent progression to HGD/EAC

To independently evaluate the association between estimated BE tissue age and subsequent neoplastic progression, we applied our BE onset age model to an independent NIBR-Progressor case-control cohort with longitudinal follow-up. This cohort included non-dysplastic BE biopsies obtained before progression to HGD or EAC in patients who subsequently developed neoplasia (“progressors”), alongside matched patients who did not progress to HGD/EAC (“non-progressors”). The initial cohort comprised 84 patients (42 progressors and 42 non-progressors). Following quality control, we estimated BE onset ages and dwell times for 66 patients with methylation data available for at least 30 BE clock CpGs, using all available clock CpGs for each patient (***Figure 6***; see **Supplementary Methods**). Across these 66 patients (n=31 progressors and n=35 non-progressors), the median estimated BE onset age was 37.6 years (IQR = 28.8-48.5), and the median estimated BE dwell time was 19.8 years (IQR = 7.3-34.7).

**Figure 6.**
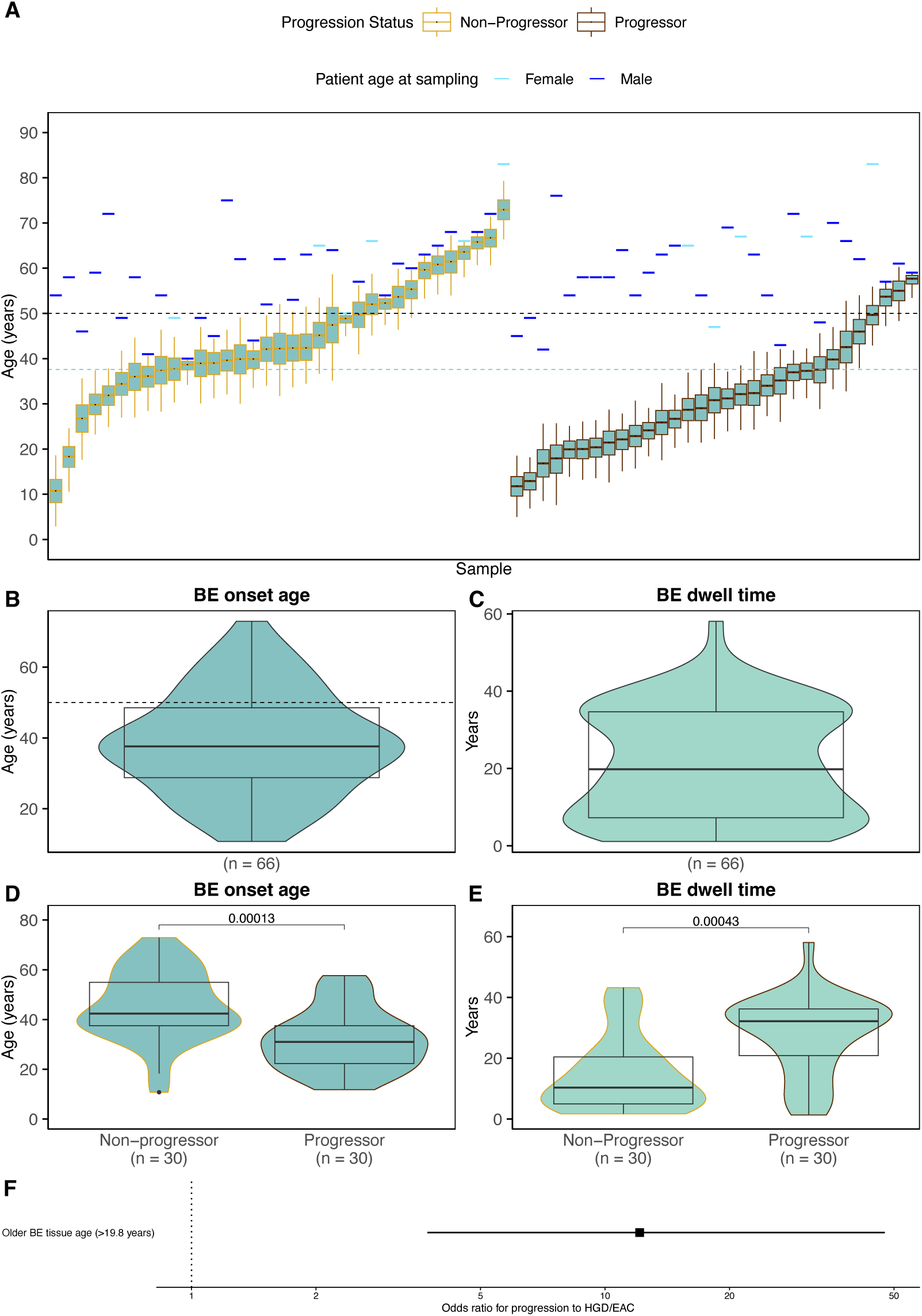
Estimated BE onset age and dwell time in patients with and without subsequent neoplastic progression. **A.** Patient ages at BE tissue sampling (horizontal bars; light blue, female; dark blue, male) and corresponding posterior distributions of estimated BE onset ages for each patient (boxplots; see **Materials and Methods**). Patients are grouped by progression status. In the overall study cohort, progressors developed HGD/EAC at least 1 year after index BE biopsy, whereas non-progressors matched for age, sex, year of BE diagnosis and follow-up time did not develop dysplasia or EAC during the corresponding follow-up period. The black horizontal dotted line indicates age 50, and the pale blue dotted line indicates the median estimated BE onset age (37.6 years) across the cohort. **B.** Violin plots comparing patient-specific median estimated BE onset ages between progressors and non-progressors. **C.** Violin plots comparing estimated BE dwell times, calculated as age at tissue sampling minus the posterior median estimated BE onset age. **D.** Paired comparison of median estimated BE onset ages between progressors and non-progressors. **E.** Paired comparison of estimated BE dwell times between progressors and non-progressors. For panels D–E, progressors and non-progressors were matched on sex and age at tissue sampling (within 2 years). **F.** Forest plot of the association between BE tissue age and subsequent HGD/EAC progression. Odds ratio (OR) and 95% confidence interval were estimated using logistic regression adjusted for chronological age at biopsy and sex. Older BE tissue age was defined as an estimated BE dwell time greater than the cohort median (19.8 years). The vertical reference line indicates OR = 1.

For these 66 patients, 24 matched pairs from the original study design were retained, and an additional six age- and sex-matched pairs were identified (age at sampling matched within 2 years), yielding 30 matched progressor/non-progressor pairs (***Supplementary Figure S8***). Among these pairs, progressors had significantly earlier estimated BE onset ages (median = 31.0 years, IQR=22.3-37.5) than non-progressors (median = 42.4 years, IQR=37.5-54.9; p=0.0001). Correspondingly, estimated BE dwell times were significantly longer in progressors (median = 32.2 years; IQR = 20.9-36.2) than in non-progressors (median = 10.4 years, IQR = 5.0-20.4; p=0.0004).

We next classified the 66 patients into younger and older BE tissue age groups using the cohort-wide median estimated dwell time of 19.8 years as the threshold (***Figure 6C***). In logistic regression analyses adjusting for chronological age at sampling and sex, patients with older BE tissue ages (>19.8 years) had significantly greater odds of subsequent neoplastic progression than those with younger tissue ages (OR = 12.1, 95% CI 3.7-47.5; p=9.96 × 10⁻⁵; ***Figure 6F***). Although the wide confidence interval reflects modest cohort size, this finding suggests that BE tissue age exceeding 20 years may represent an important threshold for surveillance risk stratification. Consistent with this finding, when BE dwell time was modeled as a continuous variable, each 14.2-year increase (equivalent to one standard deviation of BE dwell time among the 66 patients) was associated with nearly 4-fold higher odds of progression (adjusted OR = 3.94, 95% CI: 2.05-8.65).

Taken together, these findings suggest that patients undergoing BE surveillance who are similar in chronological age may nevertheless have substantially different BE tissue ages, as measured using epigenetic data derived from their bulk BE biopsy. Importantly, these differences were associated with subsequent neoplastic progression, supporting the potential utility of BE tissue age as a molecular biomarker for risk stratification beyond conventional clinical measures.

### Estimated BE onset ages supported by epidemiologic data on BE prevalence

As an independent validation of our epigenetic clock estimates, we examined published epidemiologic studies reporting age-specific BE prevalence among patients undergoing their first esophagogastroduodenoscopy (EGD)^44–46^. To compare with our primary analyses of BE onset age estimates across both sexes (***Figure 2***), we used overall BE prevalence estimates combining males and females. These studies showed remarkably consistent, approximately linear increases in BE prevalence with age (***Figure 7A***). In the largest study (n=76,691 BE patients among 2,337,786 total patients undergoing EGD), the fitted prevalence curve increased approximately linearly between ages 15 and 65 years. Under a simplified model of persistent BE, this pattern is consistent with an approximately uniform distribution of onset ages over this interval, corresponding to a mean onset age of 40 years. Consistent with age of onset distributions for other common diseases, a fitted gamma distribution also recapitulates this data when constraining the mean onset age to 40 years (***Figure 7B***, see **Supplementary Methods**). The fitted gamma distribution had a shape parameter of 6.14 and an asymptotic prevalence of 4.36%, corresponding to a median onset age of 37.9 years and a mode of 33.5 years (***Figure 7C*,** blue density).

**Figure 7.**
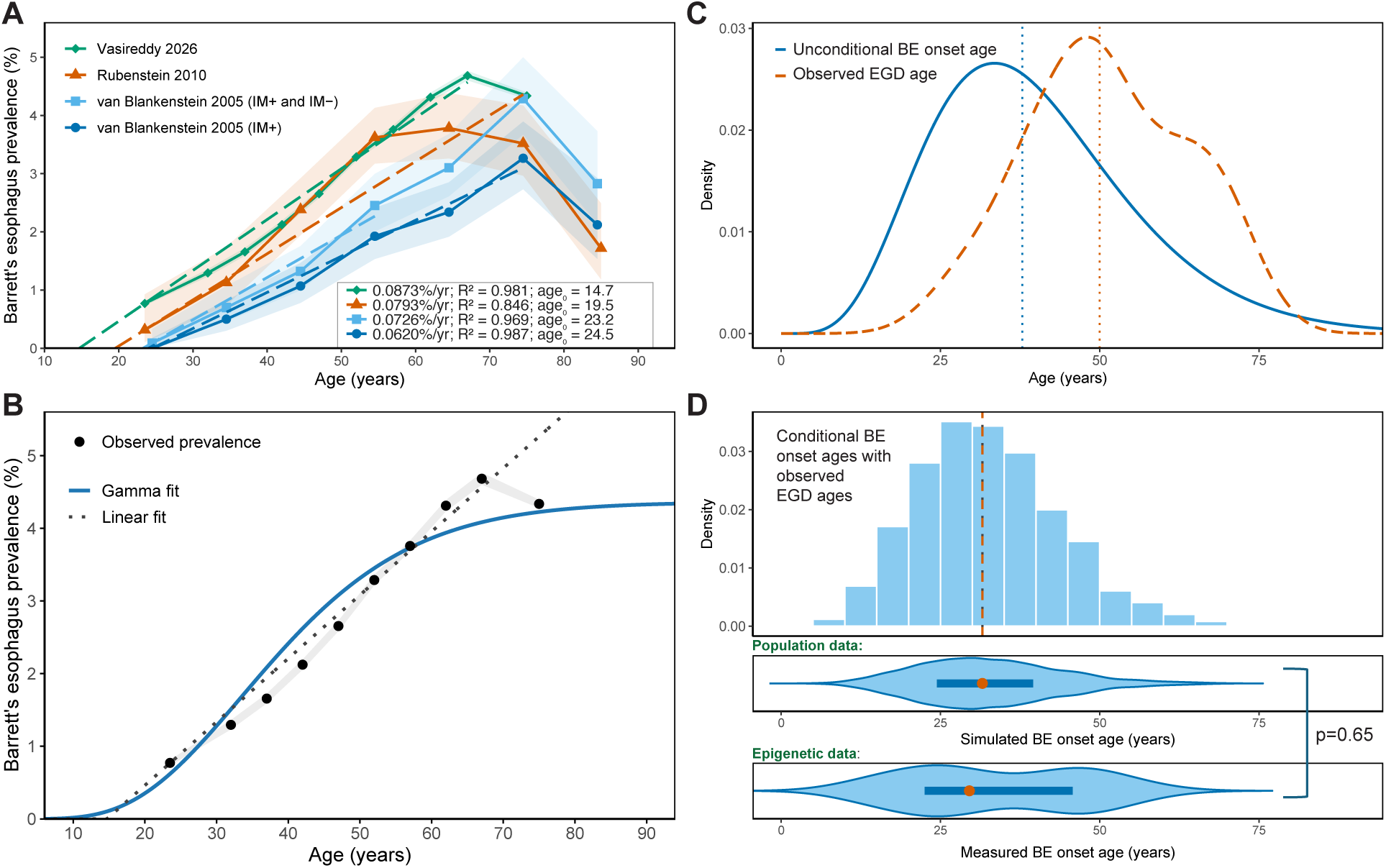
Estimated BE onset ages derived from molecular data are supported by epidemiologic data. **A.** Age-specific BE prevalence estimates from 4 epidemiologic study populations, with linear regression fits illustrating the increase in prevalence with age (x-intercept denoted by a_0_). Points indicate observed prevalence; shaded intervals represent 95% confidence intervals. One study^45^ included patients with (+) and without (-) intestinal metaplasia (IM) detected. **B.** Age-specific BE prevalence reported by Vasireddy, et al. (2026), compared with a fitted scaled gamma cumulative distribution function (solid blue line) and a linear regression model to age 65 (dashed black line). **C.** Estimated probability density of BE onset ages derived from the fitted unconditional gamma model (blue) compared with the observed distribution of ages at endoscopy (EGD) in the NIBR study cohort (orange, also shown in Figure 2B). **D.** Simulated BE onset age distribution obtained by sampling onset ages conditional on each patient’s age at EGD sampled from the NIBR cohort data (i.e., BE diagnosis ages), compared with independently estimated BE onset ages from molecular data as shown already in Figure 2B. Simulations assumed that BE onset preceded EGD.

Although this distribution is older than onset ages derived from our epigenetic data above, the distribution of onset ages among clinically ascertained prevalent cases depends on the ages at which patients are sampled^47^. Conditioning on BE being present at the time of diagnosis can therefore shift the observed onset age distribution toward younger ages. To account for this sampling effect, we simulated a cohort representative of the NIBR cohort by drawing a random sampling age (i.e., age at diagnostic EGD) with replacement from the 24 observed BE diagnosis ages (***Figure 7C*,** orange density). For each simulated patient, we then sampled a BE onset age from the fitted gamma distribution, truncated at their age of sampling (***Figure 7D***).

Among 10,000 simulated patients, the median estimated BE onset age was 31.5 years old (95% simulation-based interval for the median in cohorts of 24 patients, 26.5-36.7 years, see **Supplementary Methods**). The simulated onset age distribution was not significantly different from the distribution independently estimated from epigenetic data in the NIBR cohort (Mann-Whitney U-test p=0.65, ***Figure 7D***). These findings demonstrate that the younger BE onset ages inferred from our epigenetic clock are consistent with the underlying onset age distributions implied by epidemiologic data after accounting for age at clinical ascertainment.

## Discussion

Barrett’s esophagus (BE) is an asymptomatic precursor to esophageal adenocarcinoma (EAC) that is often undetected before cancer diagnosis. Although the annual risk of progression from non-dysplastic BE to EAC is low (0.1-0.3%), the estimated lifetime risk of ∼10%^48^ is sufficient to explain the overall burden of EAC in the US^16^. Reducing EAC mortality depends on first identifying individuals with BE and subsequently detecting and treating high-grade dysplasia (HGD) or early EAC during surveillance. Current US and European screening guidelines consider age ≥50 years as an established risk factor^10,12,23,24^, yet rising BE incidence rates rising in younger populations^25^ who have substantial prevalent neoplasia risk^46^ highlights the need to better understand when BE develops and how long it persists before clinical detection.

We applied an established BE molecular clock to DNA methylation data from 294 total samples across three UK study cohorts to estimate BE onset age and biological tissue age (dwell time). Model validation using normal gastric cardia, squamous esophagus, and gastroesophageal junction tissues demonstrated minimal epigenetic divergence from the reference normal state, compared with significantly greater dwell times in the BE tissue. Estimates were further validated using two independent epigenetic clock models.

In the population-based Northern Ireland Barrett’s Oesophagus Register (NIBR), we found the median estimated BE onset age was 29.5 years, with 87.5% of patients developing BE before age 50. Similar results were observed in the Cambridge cohort (median 29.3 years; 87.7% before age 50). In the independent NIBR-Progressor cohort, median onset age was somewhat higher (37.6 years), although 78.8% of patients still had estimated onset before age 50. To independently evaluate these estimates, we modeled BE onset using published US age-specific prevalence data. The fitted population onset-age distribution had a median of 37.9 years, compared with 31.5 years when conditioned on the endoscopy ages of the NIBR cohort, closely matching our methylation-based estimates. Simulations further demonstrated that median onset age can vary by approximately 10 years across modestly sized cohorts due to age-dependent ascertainment. These findings suggest that the population median BE onset age may be near 40 years, whereas clinically diagnosed cohorts may have younger median onset ages near 30 years. This ascertainment effect may also explain the tendency toward later estimated onset in women, who are generally diagnosed at older ages, as sex differences were not significant after matching on age at sampling.

Beyond informing BE natural history, molecular tissue age may provide a biomarker of neoplastic progression risk. Epidemiologic studies suggest that progression risk accumulates with time since BE diagnosis^27,49,50^, potentially reflecting ongoing mutation accumulation and clonal evolution. However, biological BE duration may be more informative than time since clinical diagnosis. Consistent with this hypothesis, BE dwell times were significantly longer in patients with adjacent dysplasia (median difference 14.3 years; p=0.0042) or concurrent EAC (median difference 12.5 years; p=0.00019) compared with matched patients with non-dysplastic BE. Interestingly, we previously found that patients with a family history of BE/EAC, a known risk factor for EAC progression, also had older BE tissue ages^28^, but were unable to analyze this in the current study populations. Moreover, in an age-matched nested case-control cohort, BE tissue age exceeding 19.8 years at initial diagnosis was associated with 12-fold increased odds of subsequent HGD/EAC progression. Together, these findings suggest that molecular estimates of BE duration may capture clinically relevant variation in progression risk beyond chronological patient age.

Our study has several limitations, including modest sample sizes, although findings were consistent across independent cohorts, tissue preservation methods (archival FFPE and fresh-frozen biopsies), and methylation platforms (Illumina EPIC and Twist Human Methylome Panel). Because BE onset cannot be directly observed histologically, our clock may capture an earlier molecular event preceding metaplasia rather than its precise initiation. Recent whole-genome sequencing evidence suggests that copy number alterations may arise decades before clinical BE detection^51^, supporting the possibility of early molecular evolution. Nevertheless, the agreement between methylation-based and independent epidemiologic estimates supports the biological plausibility of our inferred onset ages. Epidemiologic estimates may be affected by misclassification from diagnostic and procedure codes in the largest dataset evaluated, although similar age-specific prevalence trends were observed in two studies requiring histopathologic confirmation of BE.

Residual confounding by BE segment length, BMI, smoking, GERD, or treatment history cannot be excluded because these variables were not consistently available. Additionally, the absence of spatial sampling information in NIBR and the use of BE tissue adjacent to EAC tumors in Cambridge may introduce heterogeneity given the polyclonal nature of BE. However, our objective was to estimate the biological age of BE tissue as a potential global marker of accumulated risk, without requiring identification of the specific clone giving rise to cancer. Our findings suggest that bulk BE biopsies could provide clinically useful information, although larger longitudinal studies with spatial sampling are needed.

An additional consideration is that greater epigenetic aging in BE tissue from progressors may reflect longer dwell times, accelerated methylation drift, or both. Elevated drift rates could also indicate a broader epigenetic field effect associated with neoplasia. Although our hierarchical Bayesian model jointly estimates onset age and CpG-specific drift rates, these mechanisms cannot be fully disentangled. Nevertheless, the independent epidemiologic validation supports the plausibility of inferred onset ages, while the observed differences between progressors and non-progressors suggest that clock-derived epigenetic aging captures biologically meaningful variation in progression risk.

Overall, our findings demonstrate the potential of methylation-based molecular clocks to reconstruct BE natural history and identify patients at increased risk of neoplastic progression. More broadly, leveraging passive DNA methylation changes to estimate biological tissue age from relatively low-cost bulk methylation data may provide a broadly applicable biomarker of cancer evolution, with potential to inform risk stratification across diverse precancerous conditions and cancer types^52^. Estimating when BE develops, often decades before diagnosis, may improve computational models of BE natural history and inform future screening strategies and clinical trial design. Importantly, these findings do not support broadly lowering screening ages. Rather, the association between BE tissue age exceeding approximately 20 years and subsequent progression suggests a prolonged window for early detection and intervention. This raises the possibility that screening beginning at age 50 could identify much of the high-risk BE population before neoplastic progression, warranting further population-based modeling. The clinical priority is therefore not simply earlier BE detection but identifying those patients most likely to progress to EAC. Incorporating molecular estimates of BE duration into future risk prediction models could enable more personalized surveillance and timely intervention to prevent EAC.

## Data and code availability

Code for the BE molecular clock was published previously and is publicly available (https://github.com/yosoykit/BE_Clock_Model)^28^. De-identified epigenetic data for the NIBR and NIBR-Progressor cohorts are planned to be made available upon publication contingent on NIBR approval. De-identified data used for the Cambridge cohort have been published previously, see original publications for details^42,43^. The OCCAMS data disclosed in the publication may only be used by not-for-profit organizations and only for academic, teaching, clinical care, and educational purposes.

## Author contributions

KC and HGC conceptualized and designed the study, supervised the work, and obtained funding. TO coordinated NIBR sample retrieval, performed sample preparation, EPIC array, and QC of data, with supervision from AJM and assistance from ES and VC. RT, ES, and VC provided Twist data for the NIBR-Progressor cohort. SRR performed the bioinformatics analyses. KC and ZH performed the epidemiologic modeling analyses. DTM annotated the H&E slides. RCF, SK, GD, BN provided de-identified data for the Cambridge cohort. SRR and TO drafted the manuscript. All authors revised the manuscript. KC acted as guarantor.

## Acknowledgments

The authors would like to thank Dr. William Grady, Dr. Georg Luebeck, and Dr. John Inadomi for helpful conversations and previous methylation data from the gastroesophageal junction.

## Ethics approval and consent to participate

The Office of Research Ethics Committees Northern Ireland (ORECNI) granted ethical approval for the NIBR (Ref:15/NI/0203) to collect information without consent. Individuals have a right to opt out of registration. Ethical approval for the study of molecular age of Barrett’s esophagus was granted under approvals from the Northern Ireland Biobank (Ref: NIB19-0294, NIB21-0023). The Northern Ireland Biobank has ethical approval from ORECNI (Ref: 21/NI/0019) for the consent, collection, storage, and release of de-identified human tissue samples for research, and this approval extends to the NIB projects referenced above.

## Patient and public involvement

Patients or the public were not directly involved in the design, or conduct, or reporting, or dissemination plans of our research.

## REFERENCES

1. Coleman, H. G. et al. Increasing incidence of Barrett’s oesophagus: a population-based study. Eur. J. Epidemiol. 26, 739–745 (2011).

2. Edgren, G., Adami, H.-O., Weiderpass, E. & Nyrén, O. A global assessment of the oesophageal adenocarcinoma epidemic. Gut 62, 1406–1414 (2013).

3. Kamangar, F. et al. The global, regional, and national burden of oesophageal cancer and its attributable risk factors in 195 countries and territories, 1990-2017: a systematic analysis for the Global Burden of Disease Study 2017. Lancet Gastroenterol. Hepatol. 5, 582–597 (2020).

4. Rodriguez, G. M. et al. Trends in Epidemiology of Esophageal Cancer in the US, 1975-2018. JAMA Netw. Open 6, e2329497 (2023).

5. Bhat, S. et al. Risk of malignant progression in Barrett’s esophagus patients: results from a large population-based study. J. Natl. Cancer Inst. 103, 1049–1057 (2011).

6. Kroep, S. et al. An Accurate Cancer Incidence in Barrett’s Esophagus: A Best Estimate Using Published Data and Modeling. Gastroenterology 149, 577–585 (2015).

7. Spechler, S. J. et al. American Gastroenterological Association technical review on the management of Barrett’s esophagus. Gastroenterology 140, e18–52; quiz e13 (2011).

8. Coleman, H. G. et al. Symptoms and endoscopic features at barrett’s esophagus diagnosis: implications for neoplastic progression risk. Am. J. Gastroenterol. 109, 527–534 (2014).

9. Honing, J. & Fitzgerald, R. C. Categorizing Risks within Barrett’s Esophagus To Guide Surveillance and Interception; Suggesting a New Framework. Cancer Prev. Res. (Phila. Pa.) 16, 313–320 (2023).

10. Fitzgerald, R. C. et al. British Society of Gastroenterology guidelines on the diagnosis and management of Barrett’s oesophagus. Gut 63, 7–42 (2014).

11. Wani, S. et al. AGA Clinical Practice Guideline on Surveillance of Barrett’s Esophagus. Gastroenterology 169, 1184–1231 (2025).

12. Weusten, B. L. A. M. et al. Diagnosis and management of Barrett esophagus: European Society of Gastrointestinal Endoscopy (ESGE) Guideline. Endoscopy 55, 1124–1146 (2023).

13. Bird-Lieberman, E. L. et al. Population-based study reveals new risk-stratification biomarker panel for Barrett’s esophagus. Gastroenterology 143, 927–935.e3 (2012).

14. Vennalaganti, P. et al. Discordance Among Pathologists in the United States and Europe in Diagnosis of Low-Grade Dysplasia for Patients With Barrett’s Esophagus. Gastroenterology 152, 564–570.e4 (2017).

15. van Putten, M. et al. ‘Missed’ oesophageal adenocarcinoma and high-grade dysplasia in Barrett’s oesophagus patients: A large population-based study. *United Eur*. Gastroenterol. J. 6, 519–528 (2018).

16. Curtius, K., Rubenstein, J. H., Chak, A. & Inadomi, J. M. Computational modelling suggests that Barrett’s oesophagus may be the precursor of all oesophageal adenocarcinomas. Gut 70, 1435–1440 (2020).

17. Nowicki-Osuch, K. et al. Molecular phenotyping reveals the identity of Barrett’s esophagus and its malignant transition. Science 373, 760–767 (2021).

18. Zamani, S. A. et al. Integrated epidemiological and molecular data inform the relationship between precancer and cancer states of esophageal adenocarcinoma. Nat. Med. https://doi.org/10.1038/s41591-026-04331-8 (2026) doi:10.1038/s41591-026-04331-8.

19. El-Serag, H. B. et al. Surveillance endoscopy is associated with improved outcomes of oesophageal adenocarcinoma detected in patients with Barrett’s oesophagus. Gut 65, 1252–1260 (2016).

20. Codipilly, D. C. et al. The Effect of Endoscopic Surveillance in Patients With Barrett’s Esophagus: A Systematic Review and Meta-analysis. Gastroenterology 154, 2068–2086.e5 (2018).

21. Cummings, L. C., Kou, T. D., Schluchter, M. D., Chak, A. & Cooper, G. S. Outcomes after endoscopic versus surgical therapy for early esophageal cancers in an older population. Gastrointest. Endosc. 84, 232–240.e1 (2016).

22. Rice, T. W. et al. Refining esophageal cancer staging. J. Thorac. Cardiovasc. Surg. 125, 1103–1113 (2003).

23. Muthusamy, V. R., Wani, S., Gyawali, C. P., Komanduri, S., & CGIT Barrett’s Esophagus Consensus Conference Participants. AGA Clinical Practice Update on New Technology and Innovation for Surveillance and Screening in Barrett’s Esophagus: Expert Review. Clin. Gastroenterol. Hepatol. Off. Clin. Pract. J. Am. Gastroenterol. Assoc. 20, 2696–2706.e1 (2022).

24. Shaheen, N. J. et al. Diagnosis and Management of Barrett’s Esophagus: An Updated ACG Guideline. Am. J. Gastroenterol. 117, 559–587 (2022).

25. Yamasaki, T. et al. Barrett’s esophagus patients are becoming younger: analysis of a large United States dataset. Esophagus Off. J. Jpn. Esophageal Soc. 17, 190–196 (2020).

26. Noordzij, I. C. et al. Exploring the incidence of dysplasia or adenocarcinoma in early onset Barrett’s esophagus. Endoscopy 56, 906–912 (2024).

27. Gatenby, P., Bhattacharjee, S., Wall, C., Caygill, C. & Watson, A. Risk stratification for malignant progression in Barrett’s esophagus: Gender, age, duration and year of surveillance. World J. Gastroenterol. 22, 10592–10600 (2016).

28. Curtius, K. et al. A Molecular Clock Infers Heterogeneous Tissue Age Among Patients with Barrett’s Esophagus. PLoS Comput Biol 12, e1004919 (2016).

29. Redston, M. et al. Abnormal TP53 Predicts Risk of Progression in Patients With Barrett’s Esophagus Regardless of a Diagnosis of Dysplasia. Gastroenterology 162, 468–481 (2022).

30. Killcoyne, S. et al. Genomic copy number predicts esophageal cancer years before transformation. Nat. Med. 26, 1726–1732 (2020).

31. Luebeck, E. G. et al. Identification of a key role of widespread epigenetic drift in Barrett’s esophagus and esophageal adenocarcinoma. Clin. Epigenetics 9, 113 (2017).

32. Yu, M. et al. Subtypes of Barrett’s oesophagus and oesophageal adenocarcinoma based on genome-wide methylation analysis. Gut 68, 389–399 (2019).

33. Wu, J. W. et al. Biological age in healthy elderly predicts aging-related diseases including dementia. Sci. Rep. 11, 15929 (2021).

34. Jiang, M. et al. Accelerated biological aging elevates the risk of cardiometabolic multimorbidity and mortality. Nat. Cardiovasc. Res. 3, 332–342 (2024).

35. Mak, J. K. L. et al. Clinical biomarker-based biological aging and risk of cancer in the UK Biobank. Br. J. Cancer 129, 94–103 (2023).

36. Marioni, R. E. et al. DNA methylation age of blood predicts all-cause mortality in later life. Genome Biol. 16, 25 (2015).

37. Liu, Z. et al. A new aging measure captures morbidity and mortality risk across diverse subpopulations from NHANES IV: A cohort study. PLoS Med. 15, e1002718 (2018).

38. Shibata, D. Mutation and epigenetic molecular clocks in cancer. Carcinogenesis 32, 123–128 (2011).

39. Yu, M., Hazelton, W. D., Luebeck, G. E. & Grady, W. M. Epigenetic aging: more than just a clock when it comes to cancer. Cancer Res. 80, 367–374 (2019).

40. Jammula, S. et al. Identification of Subtypes of Barrett’s Esophagus and Esophageal Adenocarcinoma Based on DNA Methylation Profiles and Integration of Transcriptome and Genome Data. Gastroenterology 158, 1682–1697.e1 (2020).

41. Katz-Summercorn, A. C. et al. Multi-omic cross-sectional cohort study of pre-malignant Barrett’s esophagus reveals early structural variation and retrotransposon activity. Nat. Commun. 13, 1407 (2022).

42. Yang, Z. et al. Correlation of an epigenetic mitotic clock with cancer risk. Genome Biol. 17, 205 (2016).

43. Teschendorff, A. E. A comparison of epigenetic mitotic-like clocks for cancer risk prediction. Genome Med. 12, 56 (2020).

44. Rubenstein, J. H., Mattek, N. & Eisen, G. Age- and sex-specific yield of Barrett’s esophagus by endoscopy indication. Gastrointest. Endosc. 71, 21–27 (2010).

45. van Blankenstein, M., Looman, C. W. N., Johnston, B. J. & Caygill, C. P. J. Age and sex distribution of the prevalence of Barrett’s esophagus found in a primary referral endoscopy center. Am. J. Gastroenterol. 100, 568–576 (2005).

46. Vasireddy, A. R. et al. The Epidemiology of Young-Onset Barrett’s Esophagus in the United States: A National Population-Based Study Using the TriNetX Database. J. Clin. Gastroenterol. https://doi.org/10.1097/MCG.0000000000002394 (2026) doi:10.1097/MCG.0000000000002394.

47. Chen, W. J., Faraone, S. V., Orav, E. J. & Tsuang, M. T. Estimating age at onset distributions: the bias from prevalent cases and its impact on risk estimation. Genet. Epidemiol. 10, 43–59 (1993).

48. Gatenby, P. et al. Lifetime risk of esophageal adenocarcinoma in patients with Barrett’s esophagus. World J. Gastroenterol. WJG 20, 9611 (2014).

49. Bhat, S. et al. Risk of malignant progression in Barrett’s esophagus patients: results from a large population-based study. J. Natl. Cancer Inst. 103, 1049–1057 (2011).

50. Nguyen, T., Thrift, A. P., Yu, X., Duan, Z. & El-Serag, H. B. The Annual Risk of Esophageal Adenocarcinoma Does Not Decrease Over Time in Patients With Barrett’s Esophagus. Am. J. Gastroenterol. 112, 1049–1055 (2017).

51. Gabbutt, C. et al. Protracted copy number alteration evolution in Barrett’s oesophagus leading to oesophageal cancer. Preprint at 10.64898/2026.10.16.26365037 (2026).

52. Gabbutt, C. et al. Fluctuating DNA methylation tracks cancer evolution at clinical scale. Nature 645, 764–773 (2025).

