## Supplementary Information for "Molecular age of Barrett’s esophagus indicates onset often occurs decades before diagnosis"

**Table of Contents:**

**I. Supplementary Methods**
**II. Supplementary References**
**III. Supplementary Figures S1-S8**

#### I. Supplementary Methods

Patients were identified from the Northern Ireland Barrett's oesophagus Register (NIBR), the only population-based register in the UK, with complete long-term follow-up for outcomes through data linkages[1–4]. An additional cohort of patients was assembled from Cambridge, composed of samples from the Oesophageal Cancer Clinical and Molecular Stratification (OCCAMS) study, the Biomarker trial, and the BEST2 study[5,6].

##### NIBR cross-sectional study population

The NIBR is a population-based registry and includes over 24,000 patients diagnosed with incident BE since 1993 in Northern Ireland. Ethical approval for the NIBR research database and data analysis was granted by ORECNI (15/NI/0203)[7]. For this cross-sectional study, two separate patient groups were identified from the NIBR: patients with persistent non-dysplastic (indolent) BE and dysplastic BE patients who had prevalent HGD. Patients with low-grade dysplasia (LGD) but not HGD were not included in either group. For the indolent group, 50 BE patients who were initially diagnosed within the Belfast Health and Social Care Trust and had consistent non-dysplastic BE over the course of 4 consecutive endoscopic biopsies (i.e., they have not progressed to HGD or EAC) were selected, with a median of 1.9 years between each surveillance biopsy, as previously reported[8]. Each patient had formalin-fixed paraffin-embedded (FFPE) blocks containing tissue from biopsies taken during their incident BE endoscopy and at least one subsequent BE endoscopy (> 6 months after their incident BE endoscopy). For 13 of these patients, we selected blocks from an additional two surveillance endoscopy time points, allowing for a total of 4 serial biopsies in these patients.

For the dysplastic group, 13 patients who had undergone endoscopic mucosal resections for HGD or EAC that were diagnosed within 6 months of their incident BE diagnosis were selected (i.e., prevalent cases rather than incident HGD/EAC progressors). Patient selection focused on patients with long-segment BE (≥3 cm) who were matched by age and sex to the 13 non-dysplastic BE patients within the first patient group containing 4 serial endoscopic biopsies. All H&E slides from each sample were blindly assessed by an expert pathologist (DTM) to confirm the presence of only BE metaplastic tissue without dysplasia for the indolent group and HGD or EAC histology for the dysplastic group. One patient from both groups, indolent and dysplastic, had a duplicate biopsy taken at the same time point to be included in the analysis as further validation.

**Cambridge study populations**

The OCCAMS consortium has recruited over 4,000 EAC patients since its founding in 2008 from 27 centers across the United Kingdom[9]. Both EAC and adjacent BE samples were collected. For the Biomarker trial and the BEST2 study, BE and normal samples were collected. For our study, non-dysplastic BE (NDBE) samples were used for BE onset age estimation and patients were grouped by highest level of neoplasia found at time the NDBE sample was taken (including concurrent EAC). A total of 164 samples were included, and 26 patients had at least one extra biopsy taken from a healthy tissue including squamous esophagus, gastric cardia, or duodenum. Samples were snap-frozen and, before DNA extraction, underwent a stringent pathology consensus review where only samples with 70% cellularity were included[5]. Samples were also reviewed by 3 independent pathologists to ensure quality[6]. Ages for BE patients without EAC (including patients with diagnoses of NDBE, indefinite for dysplasia [IND], LGD, HGD, or intramucosal carcinoma [IMC]) were recorded at the time of endoscopic procedures for BE, which was either at or after the time of BE diagnosis (see ***Table 1*** in **Main Text**), and ages for EAC patients were recorded when the BE sample was taken (either at EAC diagnostic endoscopy or a short time later from the surgical specimen). Patients with diagnoses of NDBE (n=21) had a median follow-up time of 8.5 years (IQR: 3-10). Experimental details for processing these samples and producing Infinium MethylationEPIC BeadChip Array (Illumina) data used in this study have been published previously[5,6].

**NIBR-Progressor study population**

A nested case-control cohort of 42 incident HGD/EAC progressors and matched non-progressor pairs was also identified from the NIBR, matched on age at diagnosis, sex, and year of incident BE diagnosis (n=84 total patients included). This subset was derived from a larger study of progressor/non-progressors in whom there was a mean follow-up of 10.4 years (standard deviation [SD] 6.3 years), range 1-25.3 years. Progressors were defined as patients who progressed from non-dysplastic BE to HGD and/or EAC (cases) at least one year after their incident BE, whilst non-progressors did not progress beyond non-dysplastic BE (controls). Biopsies were obtained from the incident BE diagnosis for both progressors and non-progressors. The median year for incident BE diagnosis was 2008 (range 1996-2014) and non-progressors had not developed HGD or EAC by the end of passive follow-up through data linkage to the Northern Ireland Cancer Registry by 31st December 2021. Samples underwent blinded pathological review, as outlined above. DNA was extracted from index non-dysplastic BE FFPE biopsies and methylation profiling was performed using the Twist Human Methylome Panel.

**DNA extraction and quality assessment for NIBR samples**

Using annotations from expert pathologist (DTM) for the NIBR study samples, BE biopsies and BE biopsies adjacent to HGD and EAC endoscopic mucosal resections were annotated and dissected using a scalpel before proceeding to DNA extraction. For the dysplastic patients, we specifically dissected and extracted DNA from the surrounding BE area only. Tissue from 6 slides of 10μm of DNA per sample were pooled and DNA extraction was performed using the GeneRead

DNA FFPE Kit (Qiagen) according to manufacturer's instructions. The extracted DNA was quantified through Quant-iT™ PicoGreen™ dsDNA Assay Kit (Invitrogen) to ensure sufficient DNA for downstream methylation analysis. DNA integrity was assessed by real-time qPCR using the Infinium FFPE QC Kit (Illumina). Samples with $\Delta CT < 5$ were considered suitable for analysis by the EPIC array. For the NIBR-Progressor cohort, DNA extraction was performed using the Qiagen AllPrep kit (Qiagen, Venlo, Netherlands) with spectrophotometric QC performed on the NanoDrop (Thermo Fisher Scientific, Massachusetts, USA).

**Epigenome-wide methylation analysis for NIBR samples**

*Illumina EPICv1 Microarray: NIBR*

DNA bisulfite conversion was performed with EZ DNA Methylation Kit (Zymo Research) followed by DNA restoration process using Infinium HD FFPE DNA Restore Kit (Illumina) for the FFPE samples to overcome the high rate of degraded DNA in archived samples. The Illumina EPIC array platform for methylation analysis was used, as per manufacturer protocol, to appraise epigenome-wide methylation of more than 850,000 methylation sites with quantitative site resolution through Infinium MethylationEPIC BeadChip Kit (Illumina) and iScan. The initial quality control (QC) was assessed from .idat files generated from the iScan using BeadArray Control Reporter (BACR) Software from Illumina (Illumina 2015).

*Twist Human Methylome Panel: NIBR-Progressor*

DNA samples were prepared using the Twist Human Methylome Panel (Twist Bioscience, South San Francisco, CA, USA) and sequenced on an Illumina NovaSeq 6000 platform with paired-end 2 x 100 bp reads (Illumina, San Diego, CA, USA). The Twist Human Methylome Panel is a target-enrichment methylation sequencing assay designed to capture CpG-rich and regulatory regions across the human genome. Sequencing was performed following bisulfite-based methylation library preparation, generating paired-end FASTQ files for each sample. Raw sequencing data were processed using the nf-core/methylseq Bismark workflow, version 2.6.0, with alignment to the GRCh38 human reference genome[10]. Bismark was used for bisulfite-aware alignment and methylation extraction at cytosine loci[11], generating per-sample deduplicated methylation coverage files.

**Epigenetic data preprocessing, normalization, and quality control**

*Illumina EPICv1 Microarray*

R (version 4.4.1) and the *minfi* package (version 1.50.0) were used for additional data quality control (QC)[12]. First, samples that failed bisulfite conversion were removed. Next, samples whose cumulative log median intensity for both the methylated and unmethylated channels were less than 21 were removed, a default parameter for *minfi* QC. Then probes whose detection p-value was greater than 0.01 for any sample and whose variance across samples was zero were removed. Normal-exponential out-of-band background correction with dye-bias normalization was performed using *preproccessNoob,* Lastly, SNP-associated, cross-reactive, non-CpG, and sex chromosome probes were removed. Batch effects were corrected for using *limma* (version 3.60.6) function *removeBatchEffects*[13].

*Twist Human Methylome Panel*

Per-sample Bismark methylation coverage files were used for downstream methylation matrix generation. Each coverage file contained genomic CpG coordinates, methylation percentage, methylated read count, and unmethylated read count. Beta values were calculated for each CpG site as methylation percentage divided by 100, equivalent to methylated reads divided by the total number of methylated and unmethylated reads at that locus. A per-sample minimum coverage threshold of 10 reads was initially applied; values with coverage below this threshold were set to missing (NA). Coverage filtering thresholds were subsequently assessed and adapted as required for downstream analyses to balance CpG retention with measurement reliability.

CpG loci were merged across all samples to generate a union beta-value matrix. Loci were retained if they passed the initial 10x coverage threshold in at least one sample, resulting in a final matrix of 9,952,003 CpG loci across n=84 samples. CpG loci were annotated using coordinate-based identifiers in the format chromosome:start-end, with chromosome, start, and end positions retained. As no separate manufacturer target manifest was available, the matrix represented observed CpG loci from the processed Twist Human Methylome assay rather than a manifest-filtered target list. A corresponding coverage matrix, loci annotation table, and per-sample processing summary were also generated for downstream analysis. Quality control was assessed from the sequencing and alignment outputs, including Bismark alignment metrics, deduplication rates, methylation extraction summaries, M-bias reports, FastQC metrics, Qualimap coverage metrics, and the aggregate MultiQC report.

**BE onset age and dwell time prediction from epigenetic data**

All statistical model analyses were performed using R (version 4.4.1). The BE onset age for each sample was inferred using a Bayesian model, as detailed in Curtius et al. 2016 [14]. To initially build this model, 67 clock-CpGs were identified that had concordant incremental drift across patients and differential drift when compared to normal squamous and gastric tissue CpGs. This hierarchical model incorporates longitudinal methylomic drift rates, patient age at BE sampling, and methylation data to predict the BE onset age. Given patient age and methylation data, the epigenetic onset age clock uses the Markov Chain Monte Carlo (MCMC) algorithm with Gibbs sampling, incorporating prior distributions for onset age, methylomic drift rates, and intercepts to draw posterior estimates of onset age. The latent onset age parameter is initialized to half of the patient age at time of biopsy, while the initial slope and intercept values are initialized to the mean of the priors determined previously[14]. 100,000 cycles of MCMC were run to obtain the sampled distribution of the posterior then used as estimates of the onset age for each patient sample. With posterior estimates for onset age, the dwell time or biological tissue age was then calculated as the chronological age of the patient at time of sampling - the median BE onset age.

Due to probe loss from preprocessing, the model used to predict the onset ages of the NIBR cohort contained 49 out of the original 67 clock-CpG sites previously identified while the model for the Cambridge cohort contained 54. Onset age estimates using Illumina methylation array data were found previously to be robust to different numbers and subsets of clock-CpGs[14]. The epigenetic data for the NIBR-Progressor cohort was derived using the Twist Human Methylone Panel, which contained 57 out of the original 67 clock CpGs. While 57 CpGs overlap between the platforms themselves, poor coverage at certain CpGs on the Twist platform led to NA values for some CpGs in a sample-dependent manner. The average number of clock CpGs with data across Twist samples was 45.5 (IQR = 33.5-54). Sensitivity analyses using random numbers and combinations of clock CpG subsets were performed to quantify robustness of onset age estimation. As the number of clock CpGs increases, the variance of the median BE onset age decreases and point estimates stabilized at ~30 clock CpGs. Applying a minimum threshold of 30 clock CpGs retained 66 of 84 samples with Twist data for downstream analyses. The maximum number of available clock CpGs for each sample was used to estimate onset age. Mann-Whitney U tests and logistic regression were used to compare onset ages/dwell times between groups and calculate odds of progression.

**BE tissue age estimation using previous epigenetic clock models**

To further validate our onset age estimation model, we applied previously published epigenetic clock models, EpiTOC1 and EpiTOC2, to our data and calculated the correlation between estimates from our model. EpiTOC1 reports a proxy for mitotic division as the average beta value across 385 polycomb target CpGs that are known to be unmethylated at birth and subsequently drift toward hypermethylation throughout life[15]. EpiTOC2 builds upon EpiTOC1, using 163 of the EpiTOC1 CpGs to estimate the number of mitotic divisions that have occurred in that tissue by age of sampling[16]. These epigenetic clocks were selected for independent validation of our BE onset age model because they share the same founding principles of our model, namely utilizing CpGs that undergo epigenetic drift with age, while simultaneously having no overlapping CpG loci with our BE clock CpG set.

**BE onset age estimation from epidemiologic prevalence data**

As an orthogonal approach to using epigenetic clock models, we also estimated the expected population distribution of BE onset ages using published age-specific BE prevalence data among individuals in the U.S. receiving their first esophagogastroduodenoscopy (EGD; n=77,832 total BE patients)[17–19]. For these three datasets, we used prevalence estimates (number of BE positive cases divided by number of first-time EGDs in each age group) previously extracted from one study[18] for earlier BE modeling work[20], while corresponding prevalence estimates were obtained directly from published tables in the other two studies[17,19]. For each age group j, we extracted the number of individuals evaluated by EGD ($N_j$) and the number diagnosed with BE ($Y_j$). We included nine age groups spanning 18-69 years, over which BE prevalence increased and approached a plateau. We excluded the open-ended age group ≥70 years, in which observed BE prevalence declined, potentially reflecting age-dependent selection among patients undergoing their first EGD rather than a true decline in cumulative BE onset. Each age group was represented by its midpoint. Our objective was to use these epidemiologic data to infer the underlying population distribution of BE onset ages.

Assuming BE onset is an age-dependent event and that BE rarely regresses spontaneously once established, age-specific cumulative BE prevalence can be modeled as a scaled cumulative distribution function (CDF) of the underlying BE onset age distribution, which we sought to estimate from the epidemiologic data. Specifically, we modeled age-specific BE prevalence using a scaled gamma CDF, $P(a) = \pi F_{\Gamma}(a; k, \theta)$, where $F_{\Gamma}$ is the gamma CDF, $k$ and $\theta$ are its shape and scale parameters, and $\pi$ represents the asymptotic population prevalence of BE. Model parameters were estimated by maximizing the binomial likelihood of the observed age-specific BE counts: $Y_j \sim \text{Binomial}(N_j, P(a))$ using the Nelder-Mead optimization algorithm. The fitted gamma probability density function then provided an estimate of the population distribution of BE onset ages among individuals who develop BE.

To estimate the cohort-level BE onset age distribution, we simulated a large population with EGD ages sampled from the empirical age distribution of the NIBR study cohort. For each simulated patient, a BE onset age was independently sampled from the fitted gamma distribution, truncated at their EGD age, using inverse-transform sampling, $T_i = F_{\Gamma}^{-1}(U_i \; F_{\Gamma}(a_i; k, \theta))$, where $a_i$ is the patient's EGD age and $U_i \sim \text{Uniform}(0,1)$. The simulated BE onset ages were then pooled to estimate the expected distribution of BE onset ages among patients with an EGD age distribution matching that of our observed cohort.

To quantify uncertainty in the estimated median BE onset age for a cohort of *n* patients, we performed 10,000 Monte Carlo simulations, each comprising *n* patients. In each simulation, BE onset ages were independently sampled from the patient-specific conditional gamma distributions using the observed EGD ages of the *n* patients. The median simulated BE onset age was calculated for each replicate, producing an empirical sampling distribution of the cohort median. The 2.5th and 97.5th percentiles of these 10,000 simulated medians were used to construct a model-based 95% simulation interval for the median BE onset age in a cohort of size *n*. These intervals capture variability arising from unobserved individual onset ages conditional on the fitted population model and observed EGD ages; they do not incorporate uncertainty in the epidemiologic prevalence estimates or fitted gamma parameters. In general, the underlying onset age distribution is informed by both disease prevalence and age of ascertainment[21].

#### II. Supplementary References


1. Coleman, H. G. *et al.* Increasing incidence of Barrett's oesophagus: a population-based study. *Eur. J. Epidemiol.* **26**, 739–745 (2011).
2. Bhat, S. *et al.* Risk of malignant progression in Barrett's esophagus patients: results from a large population-based study. *J. Natl. Cancer Inst.* **103**, 1049–1057 (2011).

3. van Putten, M. *et al.* 'Missed' oesophageal adenocarcinoma and high-grade dysplasia in Barrett's oesophagus patients: A large population-based study. *United Eur. Gastroenterol. J.* **6**, 519–528 (2018).
4. Bhat, S. K. *et al.* Oesophageal adenocarcinoma and prior diagnosis of Barrett's oesophagus: a population-based study. *Gut* gutjnl-2013 (2014).
5. Jammula, S. *et al.* Identification of Subtypes of Barrett's Esophagus and Esophageal Adenocarcinoma Based on DNA Methylation Profiles and Integration of Transcriptome and Genome Data. *Gastroenterology* **158**, 1682-1697.e1 (2020).
6. Katz-Summercorn, A. C. *et al.* Multi-omic cross-sectional cohort study of pre-malignant Barrett's esophagus reveals early structural variation and retrotransposon activity. *Nat. Commun.* **13**, 1407 (2022).
7. Kearney, T. M. *et al.* Validation of the completeness and accuracy of the Northern Ireland Cancer Registry. *Cancer Epidemiol.* **39**, 401–404 (2015).
8. Kunzmann, A. T. *et al.* Does Risk of Progression from Barrett's Esophagus to Esophageal Adenocarcinoma Change Based on the Number of Non-dysplastic Endoscopies? *Dig. Dis. Sci.* **66**, 1965–1973 (2021).
9. Peters, C. J. *et al.* A decade of the Oesophageal Cancer Clinical and Molecular Stratification Consortium. *Nat. Med.* **30**, 14–16 (2024).
10. Ewels, P. A. *et al.* The nf-core framework for community-curated bioinformatics pipelines. *Nat. Biotechnol.* **38**, 276–278 (2020).
11. Krueger, F. & Andrews, S. R. Bismark: a flexible aligner and methylation caller for Bisulfite-Seq applications. *Bioinformatics* **27**, 1571–1572 (2011).
12. Aryee, M. J. *et al.* Minfi: a flexible and comprehensive Bioconductor package for the analysis of Infinium DNA methylation microarrays. *Bioinformatics* **30**, 1363–1369 (2014).
13. Ritchie, M. E. *et al.* limma powers differential expression analyses for RNA-sequencing and microarray studies. *Nucleic Acids Res.* **43**, e47 (2015).
14. Curtius, K. *et al.* A Molecular Clock Infers Heterogeneous Tissue Age Among Patients with Barrett's Esophagus. *PLoS Comput Biol* **12**, e1004919 (2016).
15. Yang, Z. *et al.* Correlation of an epigenetic mitotic clock with cancer risk. *Genome Biol.* **17**, 205 (2016).
16. Teschendorff, A. E. A comparison of epigenetic mitotic-like clocks for cancer risk prediction. *Genome Med.* **12**, 56 (2020).
17. Vasireddy, A. R. *et al.* The Epidemiology of Young-Onset Barrett's Esophagus in the United States: A National Population-Based Study Using the TriNetX Database. *J. Clin. Gastroenterol.* (2026) doi:10.1097/MCG.0000000000002394.
18. Rubenstein, J. H., Mattek, N. & Eisen, G. Age- and sex-specific yield of Barrett's esophagus by endoscopy indication. *Gastrointest. Endosc.* **71**, 21–27 (2010).
19. van Blankenstein, M., Looman, C. W. N., Johnston, B. J. & Caygill, C. P. J. Age and sex distribution of the prevalence of Barrett's esophagus found in a primary referral endoscopy center. *Am. J. Gastroenterol.* **100**, 568–576 (2005).
20. Curtius, K., Rubenstein, J. H., Chak, A. & Inadomi, J. M. Computational modelling suggests that Barrett's oesophagus may be the precursor of all oesophageal adenocarcinomas. *Gut* **70**, 1435–1440 (2020).
21. Bagiella, E. Estimating and comparing the distribution of onset of disease from prevalence case-control data. *Biometrics* **62**, 54–60 (2006).

### III. Supplementary Figures

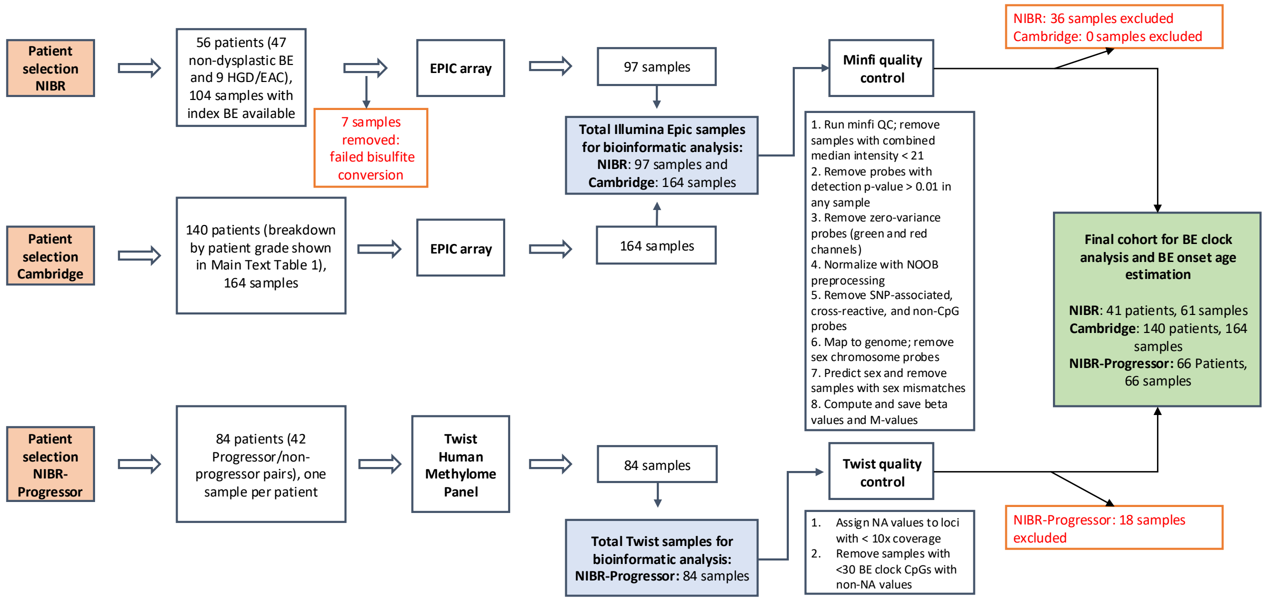


***Supplementary Figure S1.*** **Experimental design flow diagram of sample inclusion/exclusion in each methodological step for final cohort creation.**

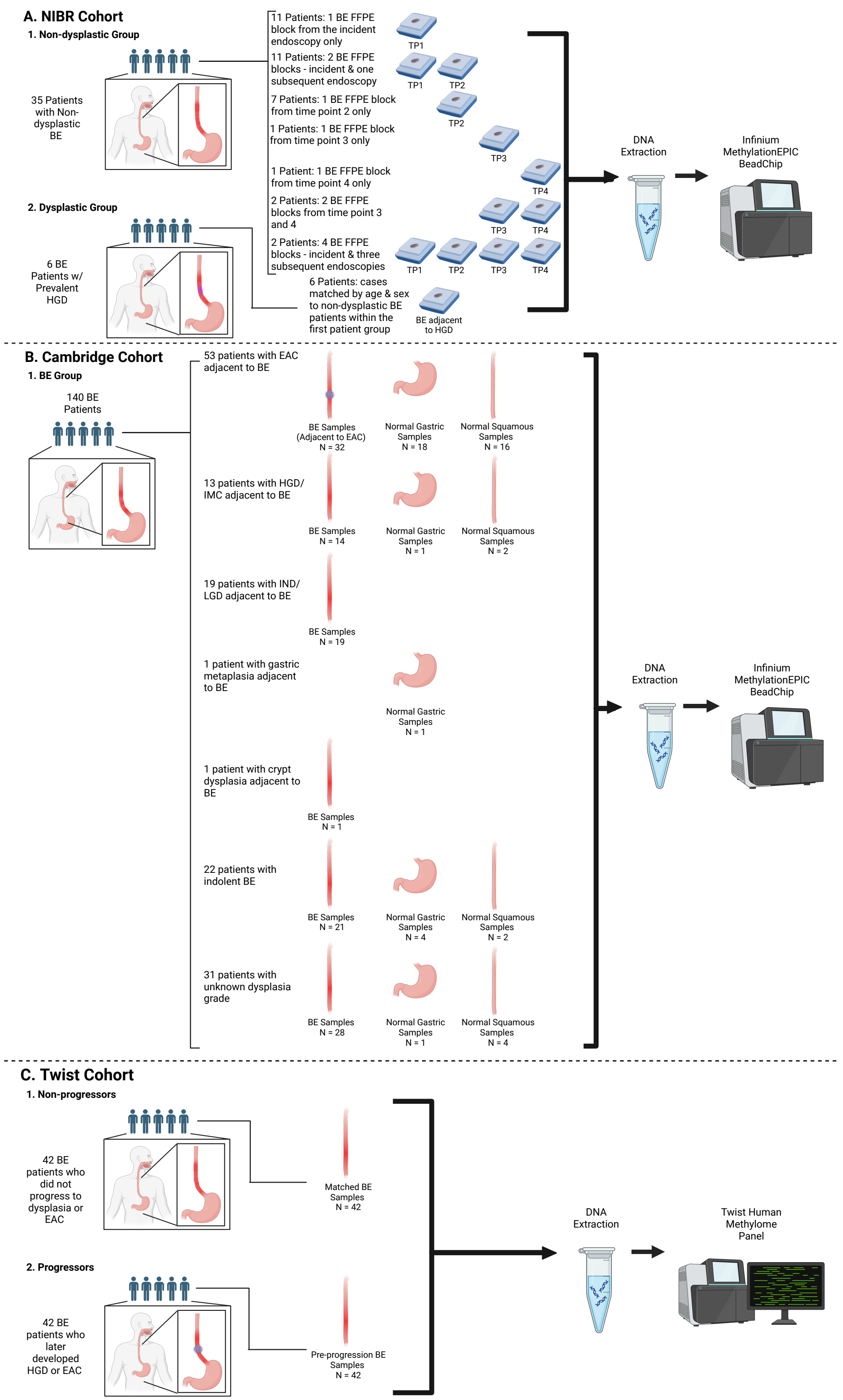


***Supplementary Figure S2.*** **Patient and sample overview in NIBR (A), Cambridge (B), and NIBR-Progressor (C) cohorts.** Sample numbers reflect those that passed initial QC filtering.

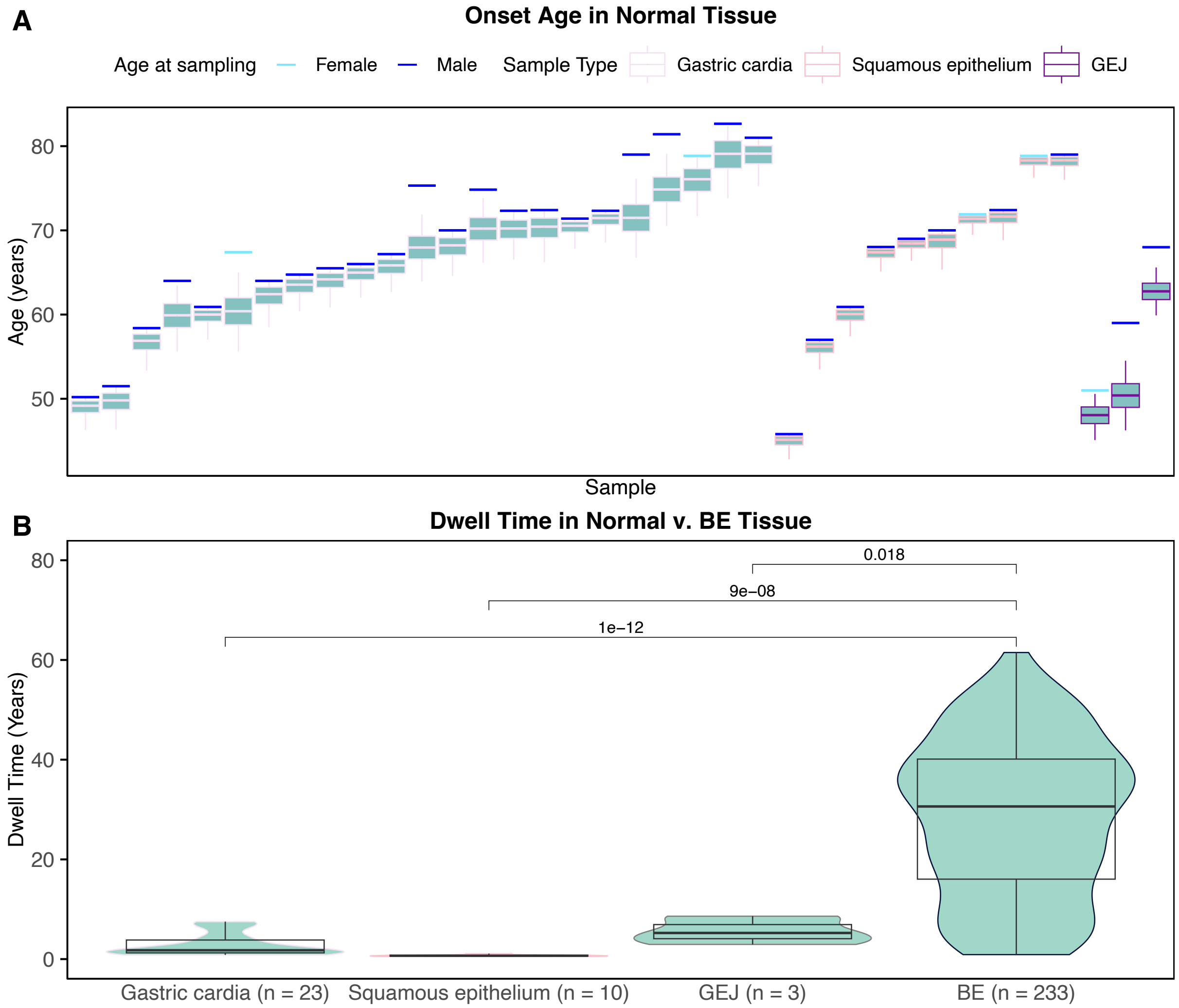


***Supplementary Figure S3.*** **BE onset age clock validation in normal gastric cardia, normal squamous esophagus, gastroesophageal junction (GEJ), and Barrett's esophagus (BE). A.** Light blue (female) and dark blue (male) horizontal bars denote age at sampling while the boxplots below show the distribution of estimated onset ages (horizontal line in box denotes median). The box length represents the interquartile range (IQR, 25th to 75th percentile) and the whiskers extend from Q1 - 1.5 x IQR to Q3 + 1.5 x IQR. **B.** Dwell time equals the age of onset subtracted from the age of sampling. All BE samples were utilized for the three cohorts (NIBR, Cambridge, NIBR-Progressor). For the two patients who had estimated BE onset ages for 2 duplicate samples, the average of their 2 duplicate samples was used.

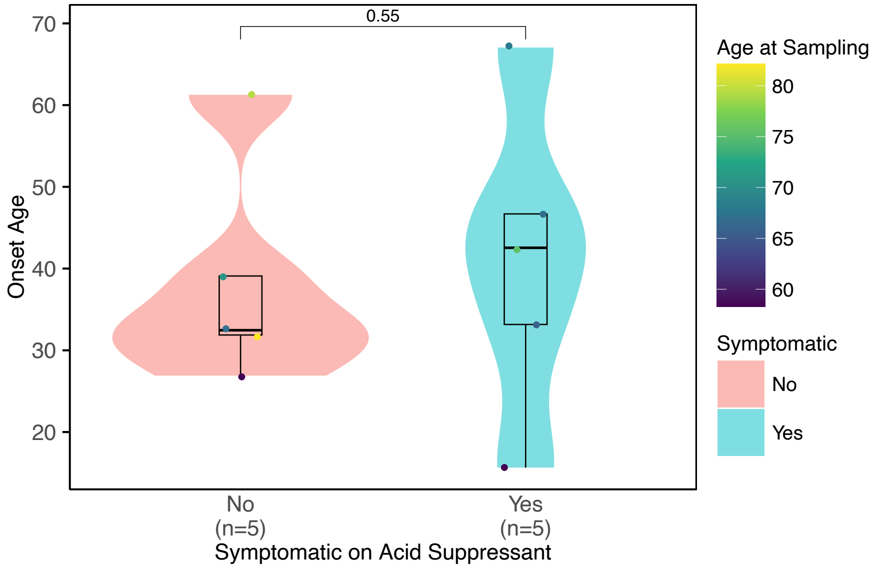


***Supplementary Figure S4*. Persistent reflux-related symptoms and estimated BE onset age.** Estimated BE onset age stratified by symptom status while receiving acid-suppressive therapy; points colored by age of patient at time of BE sampling.

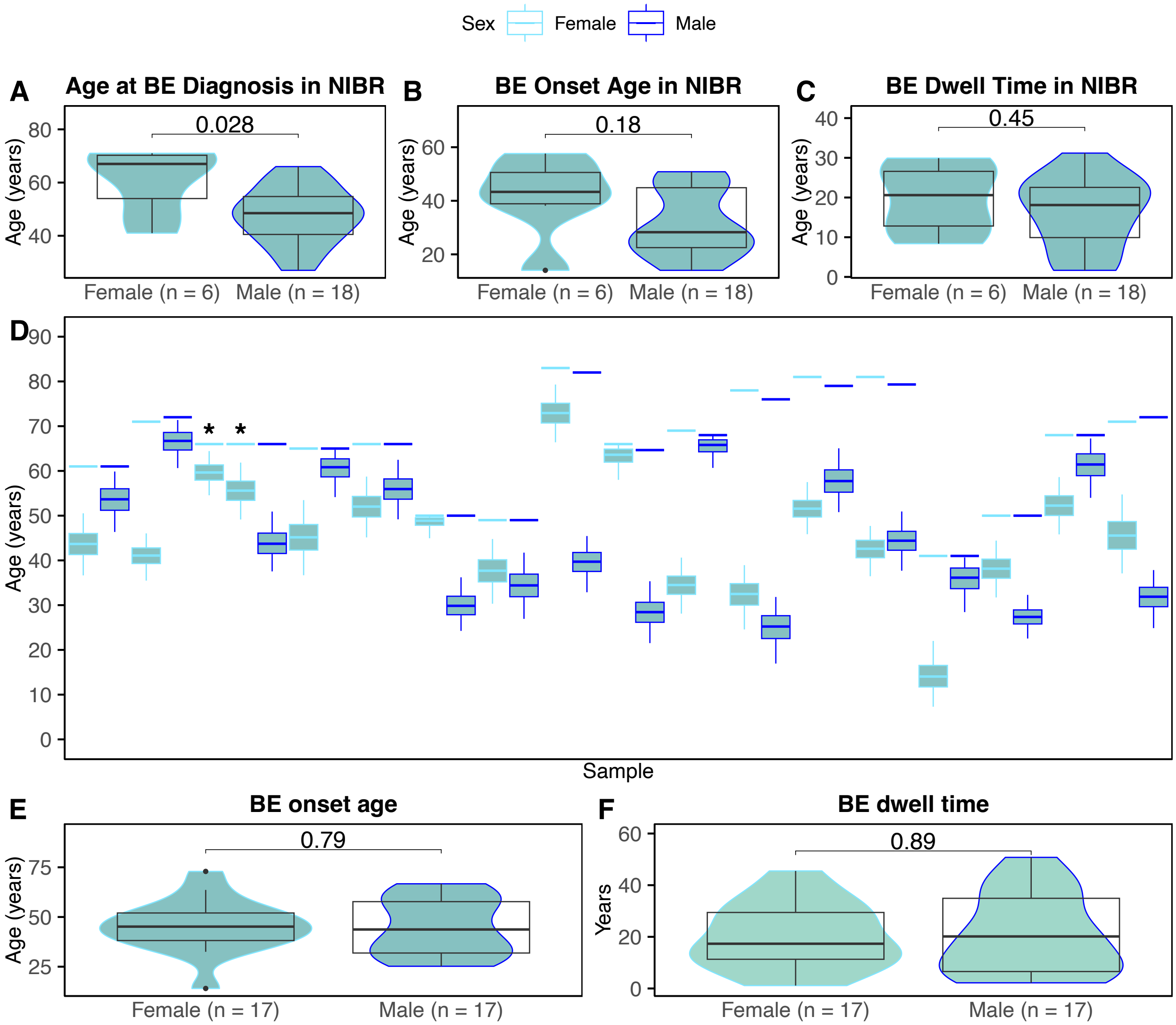


***Supplementary Figure S5*. Median BE onset age and dwell time estimates stratified by sex.** Violin plots compare distributions of each variable by sex. **A.** BE diagnosis ages in NIBR cohort. **B.** BE onset age estimates in NIBR cohort for timepoint 1 (TP1) samples taken at time of BE diagnosis. **C.** NIBR BE dwell time estimates (BE onset age subtracted from age of BE sampling). **D.** A pooled cohort was composed of TP1 samples from the NIBR cohort, samples from the Cambridge cohort that have no adjacent dysplasia (NDBE), and non-progressors from the Twist cohort. Of these, patients were matched by age (within 2 years) and sex, constituting 17 male/female patient pairs. Patient ages at BE tissue sampling (horizontal bars; light blue, female; dark blue, male) and corresponding posterior distributions of estimated BE onset ages (boxplots; see **Materials and Methods**). Average of estimated medians for duplicate samples (denoted by *) used for 1 patient. **E.** Comparison of median estimated BE onset ages between matched groups. **F.** Comparison of median estimated BE dwell times between matched groups.

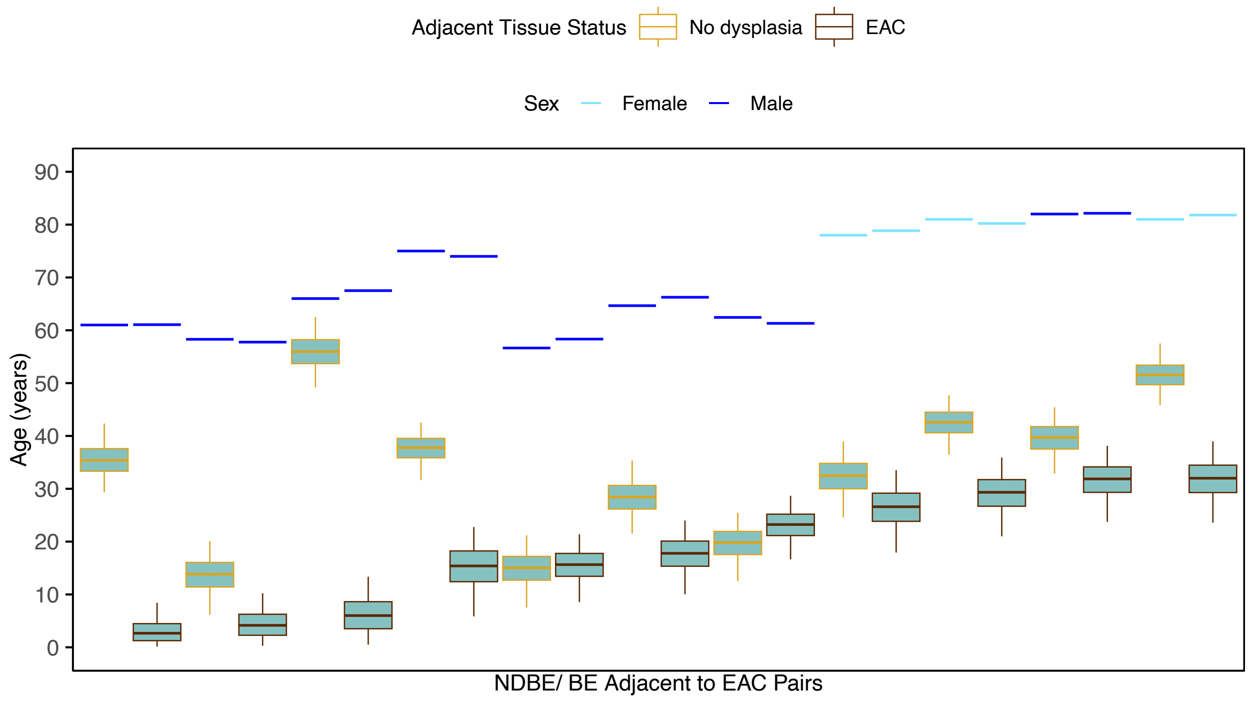


***Supplementary Figure S6.*** **Estimated BE onset ages for age- and sex-matched patients with non-dysplastic BE (NDBE) and patients with BE adjacent to EAC.** Patient ages at BE tissue sampling (horizontal bars; light blue, female; dark blue, male) and corresponding posterior distributions of estimated BE onset ages (boxplots; see **Materials and Methods**). Patients were matched by age (within 2 years) and sex.

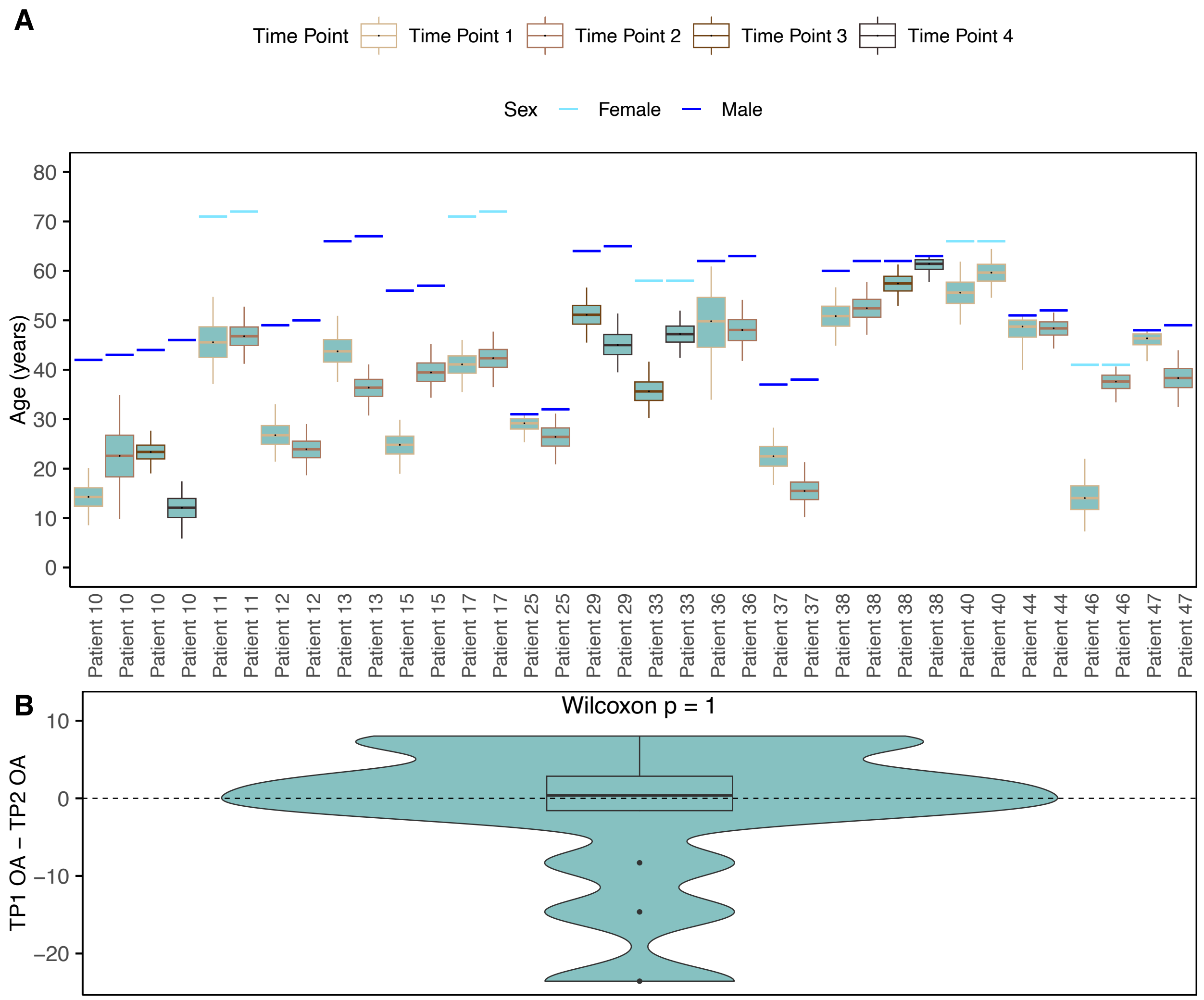


***Supplementary Figure S7*. BE onset age in comparisons in longitudinal samples with Barrett's esophagus. A.** Estimated BE onset ages for all longitudinal samples in the NIBR cohort. **B.** Per patient difference in estimated BE onset ages (OA) measured for time point 1 (i.e., time of BE diagnosis) and time point 2 (median = 0.4 years for 13 sample pairs with available data, pairwise Wilcoxon test p = 1).

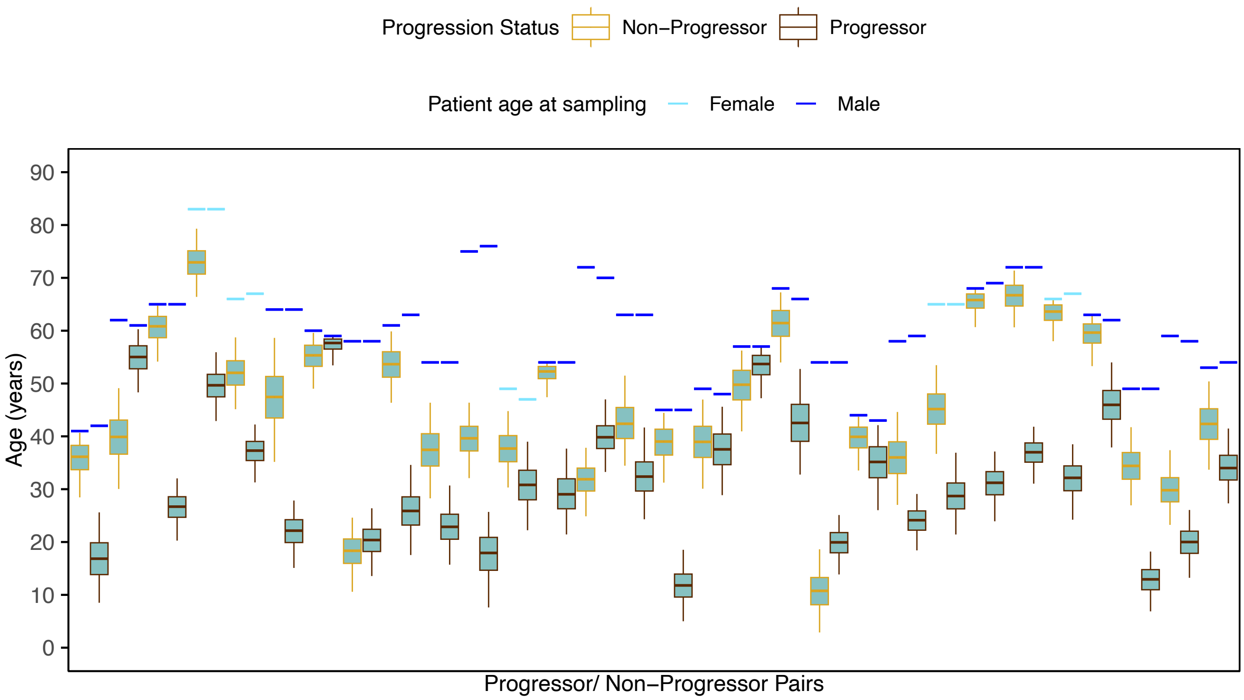


***Supplementary Figure S8.*** **Estimated BE onset ages for age- and sex-matched progressor and non-progressor patient pairs in the NIBR-Progressor cohort.** Patient ages at BE tissue sampling (horizontal bars; light blue, female; dark blue, male) and corresponding posterior distributions of estimated BE onset ages (boxplots; see **Materials and Methods**). Patients were matched by age (within 2 years) and sex.
